# Association of sedentary behavior with atrial fibrillation-related biomarkers in a population with overweight or obesity

**DOI:** 10.1101/2025.08.25.25334401

**Authors:** Linzi Li, Amit Shah, Yi-An Ko, Dayna A. Johnson, Dora Romaguera, Angel M. Alonso-Gómez, Estefanía Toledo, Cristina Razquin, Jadwiga Konieczna, Miguel Angel Martinez-Gonzalez, Julia Warnberg, Montserrat Fitó, Alvaro Alonso

## Abstract

**Background:** Blood biomarkers can characterize the atrial substrate, helping to elucidate mechanisms of atrial fibrillation (AF) development. Understanding whether sedentary behavior affects AF-related biomarkers is key for future prevention strategies.

**Methods:** We studied 252 participants in PREDIMED-Plus, a multicenter randomized trial in Spain for the primary prevention of cardiovascular disease. A wrist-worn accelerometer was used to measure physical activity (PA) in a week at baseline and at least once during follow-up at years 3 and 5. Blood samples were collected at each time point to measure selected cardiovascular biomarkers: propeptide of procollagen type I, high-sensitivity (hs) troponin T (hsTnT), hs C-reactive protein, 3-nitrotyrosine, and N-terminal propeptide of B-type natriuretic peptide (NT-proBNP). Sedentary time was assessed as inactive time (< 1.5 METs in waking time). Using isotemporal substitution, we analyzed the impact of replacing 30-min sedentary time per day with low-intensity PA (LPA, 1.5–3 METs), moderate to vigorous PA (MVPA, > 3 METs) and time in bed (time difference between going to bed and getting up) on log-transformed biomarkers cross-sectionally and longitudinally, using linear regression and mixed models.

**Results:** At baseline, 252 eligible participants averaged 65 years of age (SD 4.9), and BMI 32.2 kg/m² (SD 3.3), and 40% were female. Cross-sectionally, replacing 30-min sedentary time with LPA, MVPA, or time in bed had no significant association with biomarkers. After 5 years, replacing baseline 30-min of sedentary time per day with LPA or MVPA was associated with lower hs-TnT concentrations compared to baseline (−4%, 95% CI - 8%, −1%; −2%, 95% CI −7%, 4%, respectively). Substituting 30-min sedentary time with LPA or MVPA showed nonsignificant reductions in NT-proBNP (−3%, 95% CI −11%, 5%; - 8%, 95% CI -20%, 5%, respectively), but not a consistent association with other biomarkers.

**Conclusion:** In overweight/obese individuals, prolonged sedentary time, when compared to engaging in PA, was associated with unfavorable changes of hs-TnT over 5 years, but no significant impact on other AF-related biomarkers.

## Background

Atrial fibrillation (AF) is the most common chronic cardiac arrhythmia. The lifetime risk for AF was estimated to be 1 in 3 in persons of European ancestry, and AF increases the risk of adverse cardiac events including ischemic stroke and heart failure (HF), compounding the overall healthcare burden of AF in the population.^1–3^ However, consistently effective interventions for primary and secondary prevention of AF remain lacking, partly due to existing knowledge gaps in the pathophysiology of AF development. The underlying pathophysiology of AF is multifactorial and complex, with the onset of AF requiring a trigger that may initiate the arrhythmia by acting on a vulnerable structural and functional atrial substrate.^4,5^ Circulating biomarkers of cardiovascular risk can characterize this atrial substrate. Previous studies have reported the association between increased risk of AF and circulating biomarkers, including cardiac fibrosis (procollagen type III N-terminal propeptide [PIIINP], matrix metalloproteinases [MMP], carboxy-terminal propeptide of procollagen type I [PICP]), myocardial injury (high-sensitivity cardiac troponin T [hsTnT]), atrial stretch or overload (B-type natriuretic peptide, N-terminal pro-B-type natriuretic peptide [NT-proBNP]), and inflammation (C-reactive protein [hsCRP]). ^6–15^ Even though these biomarkers also participate in other cardiovascular disease (CVD) mechanisms and are nonspecific to AF, they can characterize the atrial substrate and help understand pathways of AF onset.

The American Heart Association’s Life’s Essential 8 recommends being active as one of the approaches to maintaining cardiovascular health.^16^ The benefits of low to moderate physical activity on both reducing the risk of AF and preventing adverse outcomes in AF patients have been frequently described.^17–20^ In addition to physical activity, existing evidence indicates that sedentary behavior is associated with an elevated risk of incident HF, stroke, total CVD, and CVD mortality.^21–23^ Oftentimes, sedentary behavior is being considered as lack of physical activity. However, recent studies recognize being sedentary as a risk factor for CVDs and CVD mortality distinct from physical activity,^24–26^ and as a correlate of CVD risk factors independent of physical activity.^27^ Using an isotemporal substitution model, the substitution of sitting with standing or stepping was found to be favorably associated with CVD risk factors.^28^ It has been suggested that the definitions of “sedentary behavior” and “physically inactive” are different, and the terminology of movement-based behavior should be consistent across studies.^29,30^

Nevertheless, few studies have explored the impact of sedentary behavior on AF development, with those studies having limitations in sedentary time assessment and evaluating highly-selected study populations.^31,32^ Moreover, the association of sedentary behaviors with the development of the AF substrate and AF-related biomarkers has not been evaluated. A better understanding of the role of sedentary behavior in the development of the AF substrate could be essential to advance our knowledge on the pathogenesis of AF and improve its management. We utilized data from PREDIMED-Plus, a randomized trial for the primary prevention of CVD in high-risk individuals, to assess the cross-sectional and longitudinal association between of sedentary behavior on blood concentrations of biomarkers of AF-related pathways, including fibrosis, myocardial damage, inflammation, oxidative stress, and atrial stretch.

## Methods

### Study design and population

The PREDIMED-Plus (Prevención con Dieta Mediterránea) study is a multicenter randomized trial in overweight and obese adults focusing on the primary prevention of CVD. Between 2013 and 2016, 6874 individuals with a body mass index (BMI) ranging from 27 to less than 40 kg/m^2^, and who met at least three criteria for the metabolic syndrome were recruited from 23 study centers across Spain. Those who had documented prevalent CVD, uncontrolled AF, active malignant cancer, or a history of malignant cancer within five years from baseline were excluded. At enrollment, participants were randomized 1:1 to an intensive lifestyle intervention (ILI) program or a control group. The ILI program is based on an energy-reduced Mediterranean diet, increased physical activity, and cognitive-behavioral weight management; the control intervention is less intensive, containing a low-intensity dietary advice on the Mediterranean diet without restricting calories (ad libitum total energy intake) only. The six-year intervention finished at the beginning of 2023, with an additional 2-year follow-up without intervention until the end of 2024. Baseline and follow-up examinations were performed during a 30-day run-in period, at randomization, at 6 months and year 1 post-randomization, and then annually. The detailed cohort profile has been described elsewhere.^33^

An ancillary study aiming to understand the effect of the PREDIMED-Plus intervention on the AF substrate was conducted in 566 participants from 3 study sites (University of Navarra, Araba University Hospital, Son Espases University Hospital). These participants had blood samples collected and underwent transthoracic echocardiography at baseline, 3 and 5 years after randomization.^33,34^ The subsample of the ancillary study showed similar characteristics as the total PREDIMED-Plus trial participants.^35^ Additionally, a subsample of 2260 participants wore an accelerometer at baseline, being invited to repeat the assessment at 6-, 12-month, and annually thereafter for at least 7 days at each time point over a 5-year follow-up period. By design, one random third of trial participants were invited to wear the accelerometer.

The trial was registered in 2014 at [www.isrctn.com/ISRCTN89898870]. The full protocol is available at [https://www.predimedplus.com/en/project/]. The ethics research committee of all study centers has approved the PREDIMED-Plus study, and this analysis was approved by the Institutional Review Board at Emory University (IRB00100921).

Among all the participants, there were 3574 men (age 55-75 years) and 3300 (age 60-75 years) women. Figure 1 shows the flowchart of the study sample in the PREDIMED-Plus trial. Among the 566 participants in the ancillary study, 13 subjects had AF at baseline and 22 had no biomarker measurements at any of the visits. Among 2260 participants who wore an accelerometer on wrist at baseline, 2189 had valid data after excluding those who had less than 3 days of data and had less than 10 hours of data each day (invalid days). Subjects that had both accelerometry data and biomarker data at baseline totaled 252. Among those participants, 98 subjects had a baseline and at least one valid follow-up on accelerometry assessments at years 3 and 5: 72 had valid data at year 3, and 65 had valid data at year 5 (Figure 1). All of the 98 subjects had follow-up information on biomarkers.

**Figure 1.**
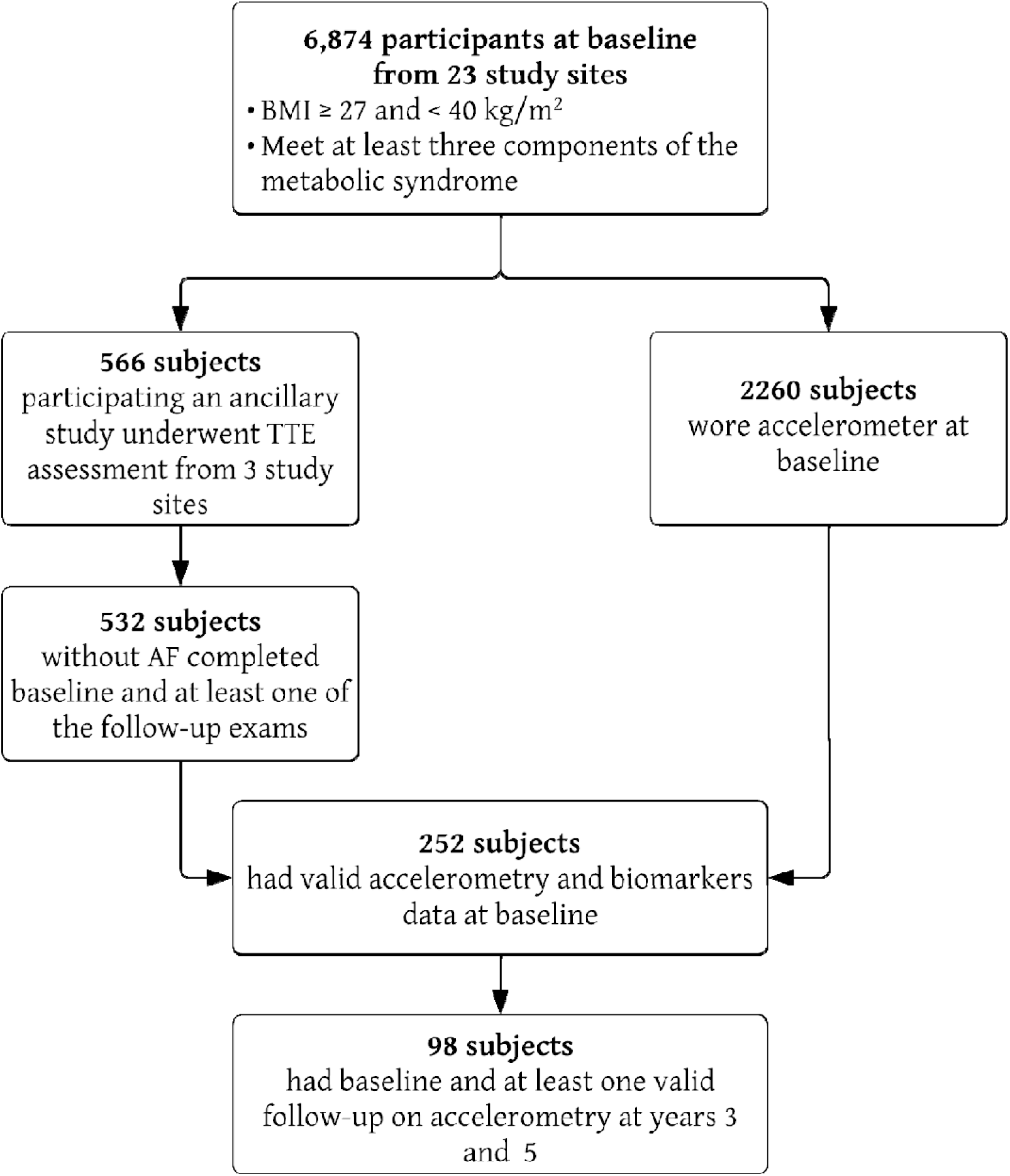
Flowchart of the study participants in PREDIMED-Plus

### Exposure measures

Sedentary behavior was measured objectively using an accelerometer (GENEActiv, ActivInsights Ltd., Kimbolton, UK). The participants were asked to wear it on their non-dominant wrist for 7 consecutive days at baseline, and at 3 and 5 years of follow-up. Sedentary time was measured using inactive time as a proxy, which was defined as any activity that requires less than 1.5 METs during waking time. Data derived from the GENEActiv device were examined as 1-min epochs of inactive time (< 1.5 METs), low-intensity physical activity (LPA, 1.5–3 METs), MVPA (> 3 METs) and time in bed (time difference between going to bed and getting up).^36,37^ Processing of raw accelerometer data has been described previously.^36^ Raw data files were managed on servers at the University of Malaga (Malaga, Spain) and processed using R software with the open-source GGIR package, version 1.2–5 (available at: cran.rproject.org/web/packages/GGIR/index.html). This open-source code has been validated through auto-calibrated functions.^38^ The autocalibration method can help reduce the calibration error in GENEActiv sensor data and improve the comparability of physical activity measures across study locations.

### Outcome measures

AF-related biomarkers concentrations were the primary outcomes in this study. The biomarkers were measured in serum samples collected at baseline and years 3 and 5 of follow-up, including carboxy-terminal propeptide of procollagen type I (PICP, marker of cardiac fibrosis), high-sensitivity troponin T (hs-TnT, marker of myocardial damage), high-sensitivity C reactive protein (hsCRP, marker of inflammation), N-terminal propeptide of B-type natriuretic peptide (NT-proBNP, marker of atrial stretch), and 3-nitrotyrosine (3-NT, marker of oxidative stress). Hs-TnT and NT-proBNP were quantified using electrochemiluminescence immunoassay (ECLIA), while hsCRP was measured via immunoturbidimetry on a Cobas 8000 autoanalyzer (Roche Diagnostics). Enzyme-linked immunosorbent assay (ELISA) was utilized to assess 3-NT and PICP with commercially available kits. Specifically, 3-NT was measured using the Human Nitrotyrosine ELISA kit (Abcam, Cambridge, UK), and PICP was analyzed with the MicroVue PICP EIA (Quidel, San Diego, CA, USA).

### Other covariates

At baseline and every yearly follow-up visit, participants completed self-reported questionnaires that collected demographic characteristics (age, sex, origin), behavioral characteristics (smoking, alcohol consumption), clinical measurements (BMI, systolic blood pressure [SBP], diastolic blood pressure [DBP], low-density lipoprotein cholesterol [LDLc], high-density lipoprotein cholesterol [HDLc]), medical status (diabetes history), and medications/hormone use (anti-hypertension, insulin, lipid-lowing medication). Clinical measurements and medical status were self-reported and checked against medical records by trained staff.

### Statistical analysis

In all analyses, biomarker concentrations were log-transformed. The baseline characteristics of the study sample were described as mean (standard deviation [SD]) for continuous variables and frequency (percentage) for categorical variables. Restricted cubic splines were used to evaluate the cross-sectional dose-response association between sedentary time and concentrations of biomarkers at baseline. We used isotemporal substitution to examine the effect of replacing 30 minutes of sedentary time with LPA, MVPA, or time in bed on the biomarkers at baseline using multiple linear regression. The models deployed the adjustments as following: 1) adjusted for sex and age; 2) further adjusted for origin, BMI, smoke status, and alcohol consumption; 3) additionally adjusted for SBP, DBP, HDL-c, LDL-c, anti-hypertension medication, lipid-lowering medication, insulin, and diabetes history. Longitudinally, the isotemporal substitution was applied in linear mixed models to evaluate: 1) the effect of substituting 30 minutes of sedentary time with LPA, MVPA, or time in bed at baseline on changes in biomarkers between baseline and years 3 and 5; 2) the effect of substituting 30 minutes of sedentary time with LPA, MVPA, or time in bed at year 3 on biomarker changes between years 3 and 5. Separate models were fit considering time as a categorical variable (baseline, year 3, year 5) or as a continuous variable (years since randomization). Random effects for both individual participants and faily units were incorporated into the mixed models, as 24 subjects had one household cohabitant also participating in the study. Covariate adjustments were identical to those in the cross-sectional analyses, with the addition of baseline accelerometer wearing time included in each model, and intervention group in the second evaluation mentioned above. Additionally, all the analyses were conducted stratifying by sex and age (<65 years and ≥ 65 years). The results were presented as percent changes, calculated as [exp(coefficient)−1]×100, along with 95% confidence intervals (CIs), based on estimates from the multivariable linear regression and mixed models. All the analyses were performed using SAS 9.4 (Cary, NC; SAS Institute Inc.).

## Results

There were 252 participants involved in this study, with an average age of 64.7 (SD 4.9) years and a BMI of 32.4 (SD 3.4) kg/m^2^. Among them, 40% were female, and 97% were of European origin. At baseline, the total sedentary time was 8.4 (SD 1.7) h/d on average. Participants spent 2.4 (SD 0.9) and 0.78 (SD 0.59) h/d engaging in LPA and MVPA. The mean time in bed was 8.1 (SD 1.2) h/d (Table 1). Over 5 years of follow-up, sedentary time slightly increased by 24 minutes per day on average (Figure 2).

**Figure 2.**
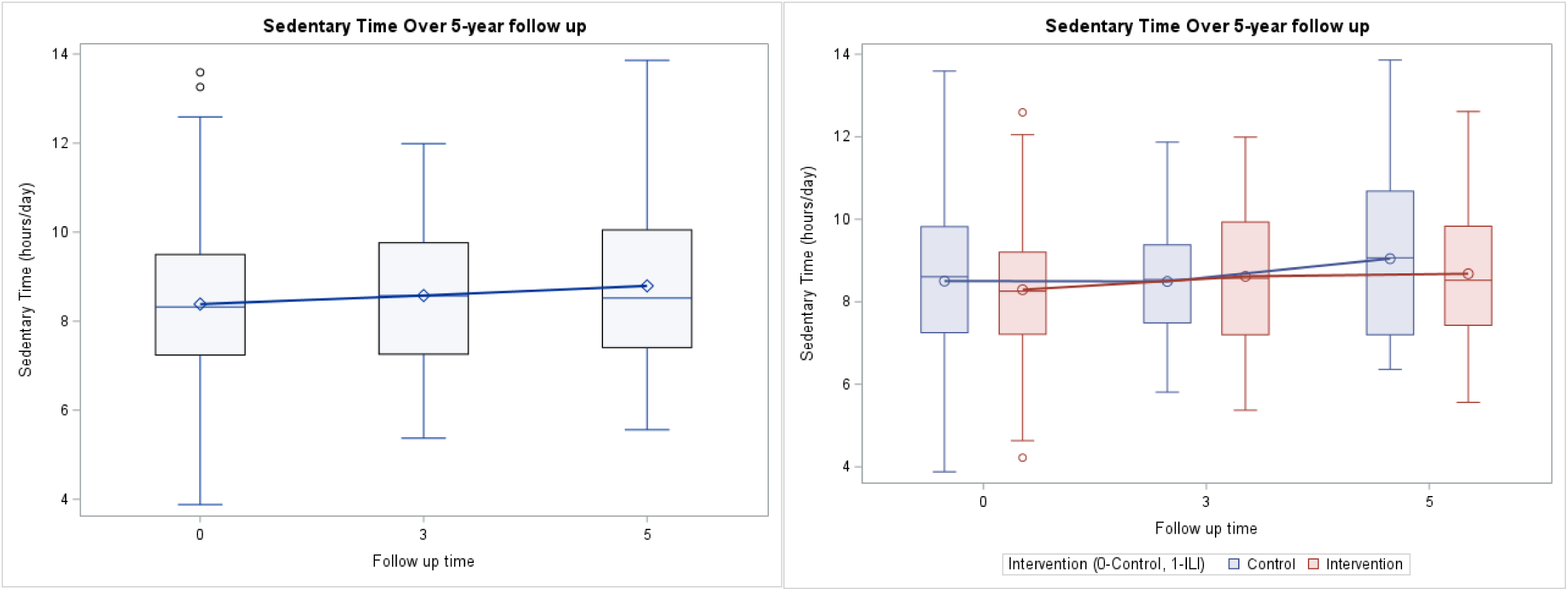
Sedentary time over 5 years follow up in the study population and by intervention groups, PREDIMED-Plus

**Table 1.** Baseline characteristics of participants in the PREDIMED-Plus trial.

|  |  |  |  |  |
| --- | --- | --- | --- | --- |
|  | Total (n=252) | Sedentary time tertiles |  |  |
|  |  | T1 (3.9-7.6 h/day) | T2 (7.7-9.1 h/day) | T3 (9.1-13.6 h/day) |
| Age, y | 64.7 (4.9) | 63.9 (4.8) | 64.6 (5) | 65.7 (4.8) |
| Female, % | 102 (40) | 34 (40) | 40 (48) | 28 (33) |
| Origin, % |  |  |  |  |
| European | 244 (97) | 82 (98) | 82 (98) | 80 (95) |
| Latin American | 8 (3) | 2 (2) | 2 (2) | 4 (5) |
| Marital status, % |  |  |  |  |
| Single | 13 (5) | 4 (5) | 6 (7) | 3 (4) |
| Married/cohabiting | 195 (77) | 64 (76) | 67 (80) | 64 (76) |
| Divorced | 25 (10) | 8 (10) | 9 (11) | 8 (10) |
| Widower | 19 (8) | 8 (10) | 2 (2) | 9 (11) |
| Education, % |  |  |  |  |
| Less than high school | 132 (52) | 46 (55) | 47 (56) | 39 (46) |
| High school | 63 (25) | 24 (29) | 16 (19) | 23 (27) |
| At least some college | 57 (23) | 14 (17) | 21 (25) | 22 (26) |
| Body-mass index, kg/m<sup>2</sup> | 32.4 (3.4) | 32 (3) | 32.4 (3.6) | 32.7 (3.5) |
| Systolic blood pressure, mmHg | 140.8 (16.9) | 138.8 (16) | 141.3 (16.7) | 142.5 (17.8) |
| Diastolic blood pressure, mmHg | 78.2 (10.5) | 78.5 (10.3) | 78.1 (11.3) | 77.9 (9.8) |
| Smoking, % |  |  |  |  |
| Never | 94 (37) | 34 (40) | 36 (43) | 24 (29) |
| Former | 133 (53) | 42 (50) | 38 (45) | 53 (63) |
| Current | 25 (10) | 8 (10) | 10 (12) | 7 (8) |
| Alcohol consumption, g/day | 14.8 (19.8) | 16.3 (22.7) | 12.4 (16.8) | 15.8 (19.3) |
| MedDiet adherence score | 7.7 (2.8) | 7.7 (2.9) | 8 (2.8) | 7.2 (2.7) |
| Diabetes, % | 76 (30) | 20 (24) | 26 (31) | 30 (36) |
| Anti-hypertension medication, % | 195 (77) | 61 (73) | 67 (80) | 67 (80) |
| Lipid-lowering medication, % | 140 (56) | 39 (46) | 49 (58) | 52 (62) |
| Insulin, % | 8 (3) | 2 (2) | 4 (5) | 2 (2) |
| Total cholesterol, mg/dL | 198.5 (32.7) | 200.6 (30.8) | 199.1 (32.8) | 195.9 (34.6) |
| Triglycerides, mg/dL | 152.5 (69.6) | 141.7 (57.8) | 150.3 (62.3) | 165.3 (84.6) |
| HDLc, mg/dL | 46.3 (11.2) | 48.1 (10.3) | 45.4 (11.7) | 45.4 (11.5) |
| LDLc, mg/dL | 122.2 (28.5) | 124.5 (26.4) | 123.9 (29.5) | 118.1 (29.5) |
| eGFR, mL/min/1.73 m<sup>2</sup> | 90.9 (12.3) | 93.3 (11.3) | 90.3 (12.3) | 89.1 (13.0) |
| PICP, mg/mL | 97.3 (40) | 96.5 (46.7) | 98.5 (37.2) | 97 (35.4) |
| hs-TnT, ng/L | 9.3 (5) | 9.1 (4.3) | 8.5 (3.8) | 10.2 (6.4) |
| hsCRP, mg/dL | 0.4 (0.6) | 0.4 (0.5) | 0.4 (0.7) | 0.4 (0.5) |
| 3-NT, nM | 728.5 (547.6) | 731.2 (525.9) | 775.2 (579) | 679 (539.1) |
| NT-proBNP, pg/mL | 73.4 (123.1) | 60.4 (50.2) | 68.1 (58) | 91.9 (198.9) |
| Time in bed, h/day | 8.1 (1.2) | 8.4 (1.2) | 8.2 (1.1) | 7.8 (1.2) |
| Sleep time excluding short awake periods, h/night | 6.3 (1.2) | 6.7 (1) | 6.2 (1.1) | 6 (1.2) |
| Sustained inactivity bouts, min/day | 1.1 (0.7) | 0.7 (0.4) | 1.1 (0.5) | 1.5 (0.8) |
| Total sedentary time, h/day | 8.4 (1.7) | 6.5 (0.9) | 8.4 (0.4) | 10.3 (1) |
| LPA, h/day | 2.4 (0.9) | 3.1 (0.9) | 2.3 (0.7) | 1.7 (0.6) |
| MVPA, h/day | 0.78 (0.59) | 0.98 (0.53) | 0.76 (0.67) | 0.58 (0.48) |

Notes: Notes: Data are shown as frequency (percentage) or mean (SD) for continuous variables of the sample.
MedDiet: Mediterranean Diet; HDLc: high-density lipoprotein cholesterol; LDLc: low-density lipoprotein cholesterol; MVPA: moderate to vigorous physical activity; TPA: total physical activity; eGFR: estimated glomerular filtration rate; PICP: C-terminal propeptide of procollagen type I; hsTnT: high sensitivity troponin T; hsCRP: high-sensitivity C reactive protein; 3-NT: 3-nitrotyrosine; NT-proBNP: N-terminal propeptide of B-type natriuretic peptide.

At baseline, replacing 30 minutes of sedentary time per day with LPA, MVPA, and time in bed had no significant association with the concentrations of PICP, hs-TnT, and 3-NT. Replacing 30 minutes of sedentary time with MVPA was associated with a lower concentration of hsCRP (−7%, 95% CI −19%, 5%) and NT-proBNP (−8%, 95% CI −18%, 2%) after adjustment for covariates; however, these associations did not reach statistical significance (Table 2).

**Table 2.** Association of isotemporal substitution of sedentary time (30 min/day) with time in bed and physical activities with biomarker at baseline, PREDIMED-Plus trial (N=252)

|  |  | LPA | MVPA | Time in bed |
| --- | --- | --- | --- | --- |
| PICP |  |  |  |  |
|  | Model 1 | -1% (-5%, 2%) | 0% (-5%, 5%) | -1% (-4%, 2%) |
|  | Model 2 | -1% (-5%, 2%) | 0% (-6%, 5%) | -1% (-4%, 2%) |
|  | Model 3 | -2% (-5%, 2%) | -2% (-7%, 4%) | -1% (-4%, 1%) |
| Hs-TnT |  |  |  |  |
|  | Model 1 | 0% (-3%, 3%) | -1% (-5%, 4%) | -1% (-4%, 1%) |
|  | Model 2 | 0% (-3%, 3%) | 0% (-4%, 5%) | -1% (-3%, 2%) |
|  | Model 3 | 1% (-2%, 4%) | 1% (-4%, 6%) | 0% (-3%, 2%) |
| hsCRP |  |  |  |  |
|  | Model 1 | -1% (-9%, 7%) | -11% (-23%, 1%) | 2% (-4%, 9%) |
|  | Model 2 | 1% (-7%, 9%) | -6% (-18%, 5%) | 4% (-2%, 10%) |
|  | Model 3 | 2% (-6%, 10%) | -7% (-19%, 5%) | 5% (-1%, 11%) |
| 3-NT |  |  |  |  |
|  | Model 1 | 0% (-7%, 7%) | 7% (-3%, 17%) | 2% (-4%, 7%) |
|  | Model 2 | 0% (-7%, 7%) | 5% (-5%, 15%) | 2% (-4%, 7%) |
|  | Model 3 | 3% (-4%, 10%) | 3% (-8%, 14%) | 2% (-4%, 7%) |
| NT-proBNP |  |  |  |  |
|  | Model 1 | -4% (-11%, 2%) | -4% (-13%, 6%) | 2% (-3%, 7%) |
|  | Model 2 | -5% (-11%, 1%) | -5% (-15%, 4%) | 1% (-4%, 6%) |
|  | Model 3 | -3% (-9%, 4%) | -8% (-18%, 2%) | 1% (-4%, 6%) |

Notes: Linear regression models were used.
Model 1 was adjusted for age, sex; model 2 adjusted for covariates in model 1 + origin, BMI, smoking status, alcohol consumption, and total wearing time; model 3 adjusted for covariates in model 2 + SBP, DBP, HDLc, LDLc, anti-hypertension medication, lipid-lowering medication, insulin, diabetes history. All biomarkers were log-transformed.

Substituting 30 minutes of baseline sedentary time per day with LPA, MVPA, and time in bed was not significantly associated with decreased hsCRP at year 3 compared to baseline. After 5 years of follow-up, reallocating baseline 30-min of sedentary time per day with LPA was associated with a lower hs-TnT concentration compared to baseline (−4%, 95% CI −8%, −1%) but not when substituted with MVPA (−2%, 95% CI −7%, 4%).

Substituting 30-min sedentary time with LPA or MVPA showed nonsignificant reductions in NT-proBNP (LPA: −3%, 95% CI −11%, 5%; MVPA: −8%, 95% CI −20%, 5%), but not a consistent impact on other biomarkers. When modeling time as a continuous variable (in years), substituting 30 minutes of sedentary time with LPA, MVPA, or time in bed showed no significant associations with biomarker concentrations (Table 3). Substituting 30 minutes of sedentary time at year 3 with LPA, MVPA, and time in bed did not show a consistent and significant impact on biomarker concentrations from years 3 to 5 (Table 4).

**Table 3.**
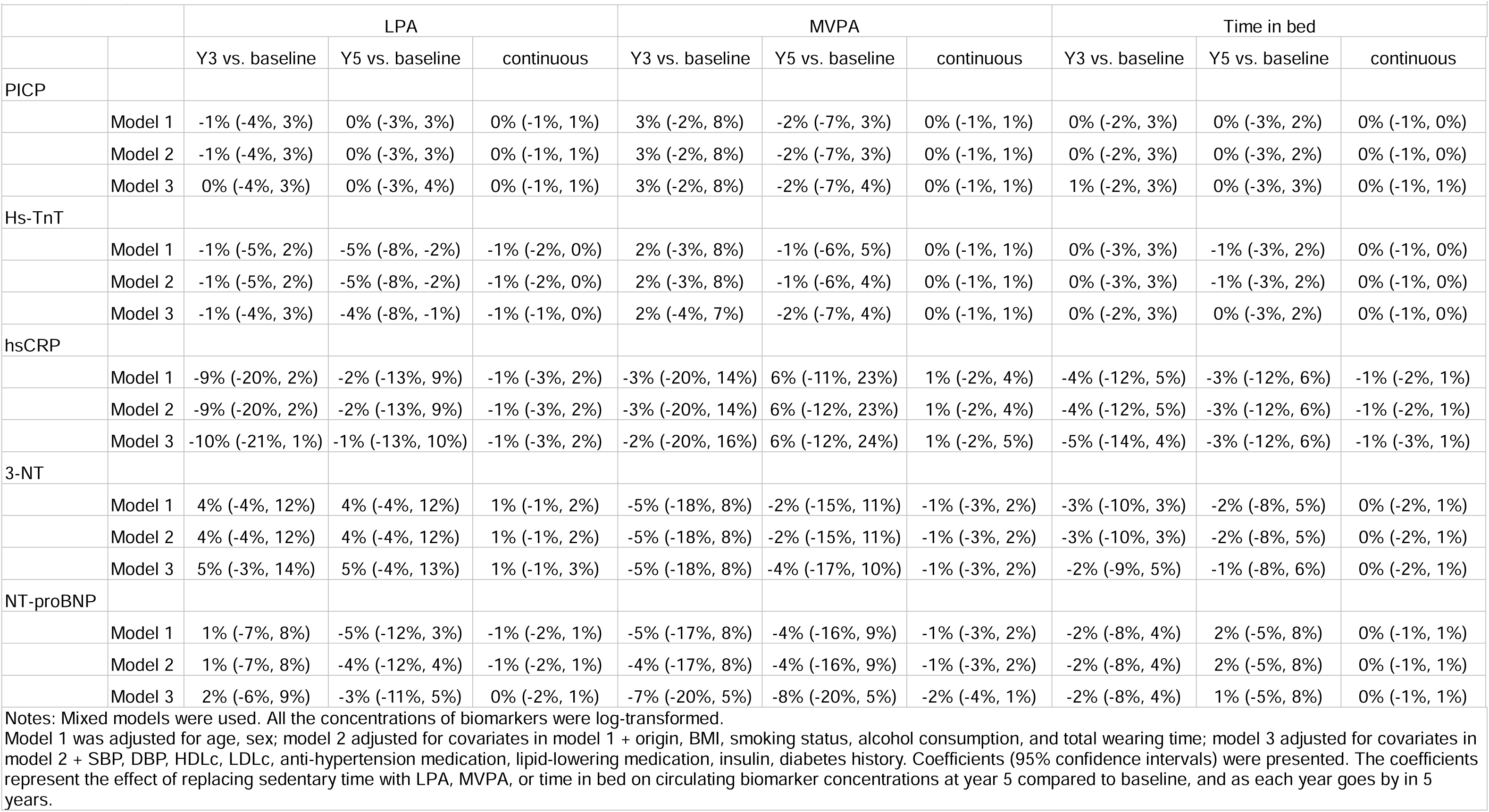
Association of isotemporal substitution of sedentary time (30 min/day) with time in bed and physical activities with changes in biomarkers between baseline and year 5, PREDIMED-Plus trail (N=98)

**Table 4.** Association of isotemporal substitution of year 3 sedentary time (30min/day) with time in bed and physical activities with changes in biomarkers between year 3 and year 5, PREDIMED-Plus trial.

|  |  | LPA |  | MVPA |  | Time in bed |  |
| --- | --- | --- | --- | --- | --- | --- | --- |
|  |  | Y5 vs. Y3 | continuous | Y5 vs. Y3 | continuous | Y5 vs. Y3 | continuous |
| PICP |  |  |  |  |  |  |  |
|  | Model 1 | 2% (-4%, 7%) | 1% (-2%, 4%) | -3% (-10%, 4%) | -2% (-5%, 2%) | -1% (-5%, 3%) | -1% (-3%, 1%) |
|  | Model 2 | 2% (-4%, 7%) | 1% (-2%, 4%) | -3% (-10%, 4%) | -2% (-5%, 2%) | -1% (-5%, 3%) | -1% (-3%, 1%) |
|  | Model 3 | 0% (-5%, 6%) | 0% (-3%, 3%) | -2% (-9%, 5%) | -1% (-5%, 3%) | -2% (-6%, 2%) | -1% (-3%, 1%) |
| Hs-TnT |  |  |  |  |  |  |  |
|  | Model 1 | -1% (-5%, 3%) | -1% (-2%, 1%) | -1% (-6%, 3%) | -1% (-3%, 2%) | 1% (-2%, 4%) | 0% (-1%, 2%) |
|  | Model 2 | -1% (-5%, 2%) | -1% (-3%, 1%) | -2% (-6%, 3%) | -1% (-3%, 2%) | 1% (-2%, 4%) | 0% (-1%, 2%) |
|  | Model 3 | -1% (-5%, 3%) | -1% (-2%, 1%) | 0% (-5%, 4%) | 0% (-3%, 2%) | 1% (-1%, 4%) | 1% (-1%, 2%) |
| hsCRP |  |  |  |  |  |  |  |
|  | Model 1 | 11% (-2%, 25%) | 6% (-1%, 12%) | 5% (-12%, 22%) | 2% (-6%, 11%) | 0% (-10%, 10%) | 0% (-5%, 5%) |
|  | Model 2 | 9% (-4%, 22%) | 5% (-2%, 11%) | 7% (-10%, 23%) | 3% (-5%, 12%) | 0% (-10%, 9%) | 0% (-5%, 5%) |
|  | Model 3 | 9% (-5%, 24%) | 5% (-3%, 12%) | 4% (-14%, 22%) | 2% (-7%, 11%) | 1% (-9%, 11%) | 1% (-5%, 6%) |
| 3-NT |  |  |  |  |  |  |  |
|  | Model 1 | -9% (-20%, 3%) | -4% (-10%, 1%) | 0% (-14%, 15%) | 0% (-7%, 7%) | -7% (-15%, 1%) | -3% (-7%, 1%) |
|  | Model 2 | -9% (-20%, 3%) | -4% (-10%, 1%) | 0% (-14%, 15%) | 0% (-7%, 8%) | -7% (-15%, 1%) | -4% (-8%, 1%) |
|  | Model 3 | -9% (-21%, 4%) | -4% (-10%, 2%) | 0% (-16%, 15%) | 0% (-8%, 7%) | -7% (-15%, 2%) | -3% (-8%, 1%) |
| NT-proBNP |  |  |  |  |  |  |  |
|  | Model 1 | 2% (-9%, 12%) | 1% (-5%, 6%) | -7% (-20%, 7%) | -3% (-10%, 4%) | -1% (-8%, 7%) | 0% (-4%, 4%) |
|  | Model 2 | 1% (-10%, 12%) | 1% (-5%, 6%) | -7% (-21%, 7%) | -3% (-10%, 4%) | -1% (-8%, 7%) | 0% (-4%, 4%) |
|  | Model 3 | 0% (-11%, 11%) | 0% (-5%, 5%) | -5% (-18%, 8%) | -3% (-9%, 4%) | 0% (-7%, 8%) | 0% (-4%, 4%) |

Notes: Mixed models were used. All the concentrations of biomarkers were log-transformed.
Model 1 was adjusted for age, sex, intervention group; model 2 adjusted for covariates in model 1 + origin, BMI, smoking status, alcohol consumption, and total wearing time; model 3 adjusted for covariates in model 2 + SBP, DBP, HDLc, LDLc, anti-hypertension medication, lipid-lowering medication, insulin, diabetes history. Coefficients (95% confidence intervals) were presented. The coefficients represent the effect of replacing sedentary time with LPA, MVPA, or time in bed on circulating biomarker concentrations at year 5 compared to year 3, and as each year goes by in 3 years.

In the stratified analysis at baseline, substituting sedentary time with LPA, MVPA or time in bed did not show an impact on biomarker levels across sex and age groups (< 65 years or ≥ 65 years at baseline) (Supplemental Table 1-2). At year 5, replacing 30-minute sedentary time per day with LPA was associated with decreased levels of hs-TnT among males (−4%, 95% CI −8%,0%) and participants aged ≥ 65 years (−3%, 95% CI −6%, 0%), but no association was observed among females (−2%, 95% CI −7%, 4%) and those < 65 years (−2%, 95% CI −5%, 1%) (Supplemental Table 3-4). Among those <65 years, replacing 30 minutes of sedentary behavior with time in bed at year 3 was related to a lower 3-NT per year from year 3 to year 5 (−8%, 95% CI −15%, 0%). However, no similar associations were found among those ≥ 65 years or when stratified by sex (Supplemental Table 5-6).

## Discussion

In this study of 252 overweight or obese participants in Spain, we found that replacing sedentary time with LPA was associated with a decreased level of hs-TnT over 5 years. Similar associations were observed for NT-proBNP without statistical significance. For PICP, hsCRP, and 3-NT, no consistent impact of substituting sedentary time was found at baseline and over time.

Hs-TnT is a well-established cardiac biomarker of subclinical myocardial damage and is used clinically to diagnose acute myocardial infarction. Previous studies have found hs-TnT to be associated with an increased risk of AF independent of other AF risk factors.^8,39^ Exercise has been reported to stimulate the temporary release of cardiac troponin, but the existing literature to date presents conflicting evidence.^40,41^ It is well-established that physical activity can improve cardiovascular health, and multiple metabolic phenomena and cardiac biomarkers are involved including hs-TnT.^42^ However, few studies have examined the impact of sedentary behavior on cardiac biomarkers, particularly those using objective measurements while accounting for physical activity. In the study Seniors-ENRICA-2, a cohort study of community-dwelling older adults in Spain, the investigators found that more PA and less sedentary behavior were associated with lower levels of hs-TnT and NT-proBNP. The associations differed by sex, subclinical damage and PA levels.^43^ In the Maastricht Study, a population study in the Netherlands, no consistent associations between sedentarism and PA with hs-TnT and high-sensitivity troponin I (hs-TnI) were observed; MVPA was found to be associated with a lower NT-proBNP level, especially if performed regularly.^44^ Among older British men, higher levels of objectively-measured sedentary behavior were linearly associated with increased levels of hs-TnT and NT-proBNP.^42^ However, the studies mentioned above utilized a cross-sectional design that limited establishing temporality between sedentary behavior and cardiac biomarkers. Furthermore, different characteristics of the study populations could also account for the inconsistency of results. In the National Health and Nutrition Examination Survey (NHANES), with accelerometer-measured PA and sedentary behavior, greater sedentary time was associated with higher CRP levels among people with short sleep; replacing 30 minutes of sedentary time with MVPA was significantly associated with reduced CRP levels.^45^ Findings from other populations have yielded mixed results regarding these associations.^46,47^

The physiology and pathophysiology of sedentary behavior on the cardiovascular system are not fully understood. Existing evidence shows that excessive and prolonged sedentary behavior is linked to a range of adverse physiological changes, including insulin resistance, vascular dysfunction, a shift in substrate utilization toward carbohydrate oxidation, decreased cardiorespiratory fitness, loss of muscle mass, strength, and bone density, as well as increases in total body fat, visceral fat accumulation, blood lipid levels, and systemic inflammation.^48^ Reducing or interrupting sedentary behavior has been found to be associated with decreased inflammation, oxidative stress, and blood pressure, and increased flow-mediated dilation.^48^ These physiological changes may influence circulating biomarker levels and, in turn, contribute to an increased risk of AF development. However, potential physiological and pathophysiological explanations about sedentary behavior and specific biomarkers are lacking. Research investigating the underlying mechanisms of sedentary behavior across diverse populations remains limited.

Our study has strengths. First, in PREDIMED-Plus, blood biomarker concentrations and actigraphy data were collected at multiple time points, including baseline and throughout the follow-up period. The repeated measurements, which offer greater insight than single-time assessments, allowed us to perform longitudinal analyses to better understand changes in biomarkers over time. Second, objectively measured sedentary behavior and PA were utilized to capture the intensity, frequency, and duration of each movement more precisely, especially for sedentary behavior, which is often misreported in self-reported data. Third, the isotemporal model provides more realistic estimates of the health benefits of replacing sedentary behavior with physical activity compared to conventional regression models. The estimates also allow for more tailored public health recommendations and policy making.

There are limitations in our study. First, the numbers of participants who had both accelerometry data and biomarker measurements are relatively small over 5 years of follow-up, resulting in imprecise estimates. Second, the accelerometer used was wrist-worn 3-axial, which cannot distinguish standing from sitting or reclining postures. Thus, there is potential for an overestimation of sedentary time due to the misclassification of standing, inactive time as sedentarism, which refers only to sitting and reclining postures; and time in bed was not equivalent to sleep time. However, time in bed was calculated using a validated algorithm unaided by sleep diary in this study.^36^ Third, the study population in PREDIMED-Plus was older, overweight or obese, and at high risk of CVD. The findings of this study may not be generalizable to populations at lower risk of CVD. Fourth, residual confounding might be an issue.

## Conclusion

In this study, among an obese and overweight population with a high risk of CVD, substituting sedentary behavior with LPA was beneficial for a more favorable level of hs-TnT over 5 years. For PICP, hsCRP, 3-NT, and NT-proBNP, no significant and consistent associations with sedentary behavior were observed. Further studies are needed in other populations to better understand the impact of sedentarism on AF development.

## Data Availability

Data collaboration for PREDIMED-Plus study is guided by the Data Sharing and Management guide. We follow a controlled data collaboration model, using anonymised (de-identified) study data only, for collaborating with approved researchers. Requests are considered by the PREDIMED-Plus Steering Committee. Please see https://www.predimedplus.com/en/project/#top for detail.

## Acknowledgment

The authors would like to thank the colleagues, staff, and participants of the PREDIMED-Plus study for their important contributions.

