## Supplementary Materials for "Association of sedentary behavior with atrial fibrillation-related biomarkers in a population with overweight or obesity"

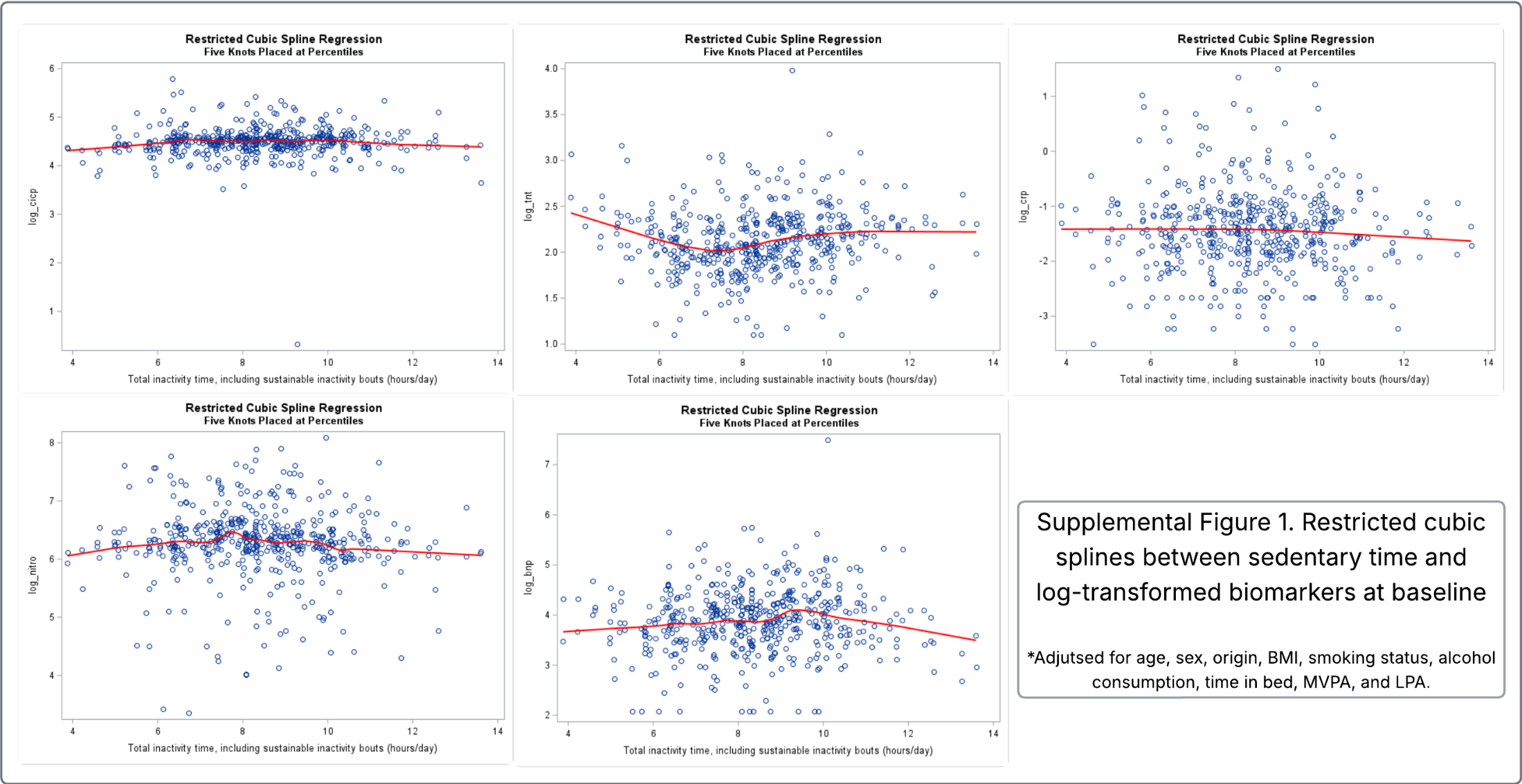

| Supplemental Table 1. Association of isotemporal substitution of sedentary time (30min/day) with time in bed and physical activities with biomarker concentrations at baseline by sex | | | | | | | | | | |
| --- | --- | --- | --- | --- | --- | --- | --- | --- | --- | --- |
|  |  | Male (n=150) | | | Female (n=102) | | | P value for interaction | | |
|  |  | LPA | MVPA | Time in bed | LPA | MVPA | Time in bed | LPA | MVPA | Time in bed |
| PICP |  |  |  |  |  |  |  |  |  |  |
|  | Model 1 | -3% (-7%, 1%) | -1% (-6%, 3%) | -1% (-4%, 2%) | 3% (-5%, 11%) | 5% (-8%, 19%) | -1% (-7%, 4%) | 0.12 | 0.37 | 0.99 |
|  | Model 2 | -3% (-7%, 1%) | -2% (-7%, 3%) | -1% (-4%, 2%) | 3% (-5%, 11%) | 5% (-10%, 19%) | -1% (-7%, 4%) | 0.11 | 0.38 | 0.94 |
|  | Model 3 | -4% (-8%, 0%) | -3% (-8%, 3%) | -1% (-4%, 3%) | 4% (-5%, 12%) | 4% (-12%, 19%) | -1% (-7%, 4%) | 0.07 | 0.29 | 0.94 |
| Hs-TnT |  |  |  |  |  |  |  |  |  |  |
|  | Model 1 | -1% (-5%, 4%) | 0% (-6%, 6%) | 0% (-3%, 4%) | 0% (-5%, 5%) | -4% (-13%, 6%) | -3% (-6%, 1%) | 0.91 | 0.54 | 0.18 |
|  | Model 2 | 0% (-4%, 4%) | 0% (-5%, 6%) | 1% (-3%, 4%) | 0% (-6%, 5%) | -2% (-12%, 8%) | -2% (-6%, 1%) | 0.8 | 0.75 | 0.15 |
|  | Model 3 | 0% (-4%, 4%) | 2% (-4%, 8%) | 1% (-3%, 4%) | 1% (-5%, 7%) | -1% (-12%, 9%) | -2% (-6%, 2%) | 0.79 | 0.85 | 0.25 |
| hsCRP |  |  |  |  |  |  |  |  |  |  |
|  | Model 1 | -3% (-14%, 8%) | -11% (-25%, 3%) | 2% (-7%, 10%) | 4% (-9%, 17%) | -15% (-38%, 8%) | 5% (-4%, 14%) | 0.23 | 0.95 | 0.90 |
|  | Model 2 | -1% (-11%, 10%) | -10% (-24%, 5%) | 4% (-5%, 13%) | 4% (-8%, 16%) | -5% (-27%, 17%) | 5% (-3%, 14%) | 0.32 | 0.96 | 0.99 |
|  | Model 3 | 0% (-12%, 11%) | -13% (-28%, 3%) | 6% (-3%, 15%) | 8% (-5%, 20%) | -6% (-30%, 18%) | 6% (-2%, 15%) | 0.30 | 0.84 | 0.84 |
| 3-NT |  |  |  |  |  |  |  |  |  |  |
|  | Model 1 | -2% (-12%, 7%) | 7% (-5%, 19%) | 3% (-5%, 10%) | 6% (-5%, 17%) | 6% (-15%, 27%) | 1% (-7%, 8%) | 0.26 | 0.71 | 0.84 |
|  | Model 2 | -2% (-12%, 7%) | 5% (-7%, 18%) | 3% (-5%, 10%) | 6% (-6%, 18%) | 4% (-18%, 25%) | 1% (-7%, 9%) | 0.28 | 0.59 | 0.91 |
|  | Model 3 | 1% (-9%, 11%) | 2% (-12%, 16%) | 3% (-5%, 11%) | 8% (-4%, 20%) | -1% (-24%, 22%) | 0% (-8%, 8%) | 0.42 | 0.73 | 0.65 |
| NT-proBNP |  |  |  |  |  |  |  |  |  |  |
|  | Model 1 | -6% (-14%, 2%) | 3% (-8%, 13%) | 3% (-4%, 9%) | -4% (-15%, 8%) | -24% (-45%, -3%) | 1% (-7%, 9%) | 0.8 | 0.02 | 0.85 |
|  | Model 2 | -7% (-15%, 1%) | 2% (-9%, 13%) | 2% (-5%, 9%) | -5% (-16%, 7%) | -28% (-50%, -7%) | 0% (-8%, 8%) | 0.87 | 0.01 | 0.78 |
|  | Model 3 | -4% (-13%, 4%) | -2% (-14%, 10%) | 2% (-5%, 9%) | -1% (-12%, 11%) | -28% (-50%, -5%) | 1% (-7%, 9%) | 0.92 | 0.02 | 0.65 |
| Notes: Linear regression models were used.  Model 1 was adjusted for age, sex; model 2 adjusted for covariates in model 1 + origin, BMI, smoking status, alcohol consumption, and total wearing time; model 3 adjusted for covariates in model 2 + SBP, DBP, HDLc, LDLc, anti-hypertension medication, lipid-lowering medication, insulin, diabetes history. All biomarkers were log-transformed. | | | | | | | | | | |

| Supplemental Table 2. Association of isotemporal substitution of sedentary time (30min/day) with time in bed and physical activities with biomarker concentrations at baseline by age | | | | | | | | | | |
| --- | --- | --- | --- | --- | --- | --- | --- | --- | --- | --- |
|  |  | <65 years (n=124) | | | >=65 years (n=128) | | | P value for interaction | | |
|  |  | LPA | MVPA | Time in bed | LPA | MVPA | Time in bed | LPA | MVPA | Time in bed |
| PICP |  |  |  |  |  |  |  |  |  |  |
|  | Model 1 | -4% (-8%, 0%) | -1% (-6%, 5%) | 0% (-3%, 3%) | 2% (-4%, 7%) | 1% (-9%, 11%) | -3% (-8%, 2%) | 0.17 | 0.69 | 0.4 |
|  | Model 2 | -4% (-8%, 0%) | -2% (-8%, 4%) | 0% (-3%, 3%) | 1% (-5%, 7%) | 2% (-9%, 12%) | -3% (-8%, 2%) | 0.19 | 0.59 | 0.47 |
|  | Model 3 | -5% (-10%, 0%) | -2% (-8%, 5%) | 0% (-4%, 4%) | 0% (-6%, 7%) | 1% (-10%, 13%) | -4% (-9%, 2%) | 0.28 | 0.48 | 0.4 |
| Hs-TnT |  |  |  |  |  |  |  |  |  |  |
|  | Model 1 | 5% (0%, 9%) | 1% (-5%, 7%) | -1% (-4%, 2%) | -5% (-10%, 0%) | -5% (-13%, 3%) | 0% (-4%, 4%) | 0.02 | 0.2 | 0.45 |
|  | Model 2 | 5% (0%, 9%) | 2% (-5%, 8%) | -1% (-4%, 3%) | -5% (-9%, 0%) | -2% (-10%, 6%) | 0% (-4%, 4%) | 0.01 | 0.36 | 0.31 |
|  | Model 3 | 6% (1%, 10%) | 1% (-5%, 8%) | -1% (-5%, 2%) | -3% (-8%, 2%) | 1% (-7%, 9%) | 1% (-3%, 4%) | 0.01 | 0.45 | 0.43 |
| hsCRP |  |  |  |  |  |  |  |  |  |  |
|  | Model 1 | 3% (-8%, 14%) | -8% (-23%, 6%) | 3% (-5%, 11%) | -4% (-16%, 7%) | -20% (-40%, 0%) | 4% (-5%, 14%) | 0.51 | 0.39 | 0.48 |
|  | Model 2 | 6% (-5%, 16%) | -3% (-18%, 12%) | 4% (-4%, 12%) | -2% (-14%, 9%) | -12% (-32%, 7%) | 6% (-3%, 15%) | 0.64 | 0.49 | 0.51 |
|  | Model 3 | 5% (-6%, 17%) | -5% (-21%, 11%) | 4% (-5%, 13%) | -1% (-13%, 11%) | -11% (-32%, 9%) | 6% (-4%, 16%) | 0.8 | 0.66 | 0.65 |
| 3-NT |  |  |  |  |  |  |  |  |  |  |
|  | Model 1 | 5% (-5%, 15%) | 2% (-12%, 15%) | 0% (-7%, 8%) | -5% (-14%, 5%) | 18% (2%, 35%) | 1% (-7%, 9%) | 0.29 | 0.25 | 0.75 |
|  | Model 2 | 4% (-6%, 14%) | -1% (-16%, 13%) | 0% (-8%, 8%) | -5% (-15%, 5%) | 19% (1%, 36%) | 1% (-7%, 9%) | 0.33 | 0.25 | 0.58 |
|  | Model 3 | 8% (-4%, 19%) | -4% (-19%, 11%) | -1% (-9%, 8%) | -3% (-14%, 8%) | 16% (-3%, 34%) | 2% (-7%, 10%) | 0.29 | 0.24 | 0.48 |
| NT-proBNP |  |  |  |  |  |  |  |  |  |  |
|  | Model 1 | 0% (-9%, 9%) | -1% (-14%, 11%) | 5% (-2%, 12%) | -9% (-18%, 0%) | -6% (-21%, 9%) | -2% (-9%, 5%) | 0.13 | 0.55 | 0.12 |
|  | Model 2 | -1% (-11%, 8%) | 0% (-14%, 13%) | 3% (-4%, 10%) | -10% (-19%, -1%) | -6% (-22%, 9%) | -3% (-10%, 5%) | 0.12 | 0.41 | 0.13 |
|  | Model 3 | 3% (-6%, 13%) | -4% (-18%, 9%) | -1% (-8%, 7%) | -9% (-18%, 0%) | -5% (-21%, 11%) | -2% (-9%, 6%) | 0.08 | 0.49 | 0.38 |
| Notes: Linear regression models were used.  Model 1 was adjusted for age, sex; model 2 adjusted for covariates in model 1 + origin, BMI, smoking status, alcohol consumption, and total wearing time; model 3 adjusted for covariates in model 2 + SBP, DBP, HDLc, LDLc, anti-hypertension medication, lipid-lowering medication, insulin, diabetes history. All biomarkers were log-transformed. | | | | | | | | | | |

| Supplemental Table 3. Association of isotemporal substitution of sedentary time (30min/day) with time in bed and physical activities with change in biomarkers between baseline and year 5 by sex | | | | | | | | | | |
| --- | --- | --- | --- | --- | --- | --- | --- | --- | --- | --- |
|  |  | LPA | | | MVPA | | | Time in bed | | |
|  |  | Y3 vs. baseline | Y5 vs. baseline | continuous | Y3 vs. baseline | Y5 vs. baseline | continuous | Y3 vs. baseline | Y5 vs. baseline | continuous |
| Male | | | | | | | | | | |
| PICP |  |  |  |  |  |  |  |  |  |  |
|  | Model 1 | 1% (-3%, 5%) | -1% (-6%, 4%) | 0% (-1%, 1%) | 0% (-7%, 7%) | 1% (-5%, 8%) | 0% (-1%, 1%) | 0% (-4%, 4%) | -4% (-8%, 0%) | -1% (-1%, 0%) |
|  | Model 2 | 1% (-3%, 5%) | -1% (-6%, 4%) | 0% (-1%, 1%) | 0% (-7%, 7%) | 1% (-6%, 8%) | 0% (-1%, 1%) | 0% (-4%, 4%) | -4% (-9%, 0%) | -1% (-1%, 0%) |
|  | Model 3 | 1% (-3%, 5%) | -1% (-7%, 4%) | 0% (-1%, 1%) | 0% (-7%, 8%) | 2% (-6%, 9%) | 0% (-1%, 2%) | 0% (-4%, 4%) | -4% (-9%, 0%) | -1% (-1%, 0%) |
| Hs-TnT |  |  |  |  |  |  |  |  |  |  |
|  | Model 1 | -1% (-4%, 2%) | -6% (-10%, -1%) | -1% (-2%, 0%) | 2% (-4%, 8%) | -1% (-7%, 5%) | 0% (-1%, 1%) | 1% (-2%, 4%) | -2% (-5%, 2%) | 0% (-1%, 1%) |
|  | Model 2 | -1% (-4%, 2%) | -6% (-10%, -1%) | -1% (-2%, 0%) | 2% (-4%, 8%) | -1% (-7%, 5%) | 0% (-1%, 1%) | 1% (-2%, 4%) | -2% (-5%, 2%) | 0% (-1%, 1%) |
|  | Model 3 | 0% (-3%, 3%) | -4% (-8%, 0%) | 0% (-1%, 0%) | 0% (-6%, 6%) | -3% (-8%, 3%) | 0% (-2%, 1%) | 1% (-2%, 4%) | -1% (-5%, 2%) | 0% (-1%, 1%) |
| hsCRP |  |  |  |  |  |  |  |  |  |  |
|  | Model 1 | -7% (-21%, 6%) | 1% (-17%, 19%) | -1% (-4%, 2%) | -13% (-37%, 11%) | 4% (-19%, 27%) | 0% (-5%, 4%) | 2% (-11%, 16%) | -1% (-16%, 13%) | 0% (-3%, 2%) |
|  | Model 2 | -6% (-20%, 7%) | 2% (-16%, 20%) | -1% (-4%, 2%) | -13% (-37%, 11%) | 3% (-20%, 27%) | 0% (-5%, 4%) | 2% (-12%, 15%) | -3% (-18%, 11%) | -1% (-3%, 2%) |
|  | Model 3 | -8% (-22%, 6%) | 5% (-14%, 24%) | -1% (-4%, 3%) | -14% (-39%, 11%) | 2% (-22%, 27%) | 0% (-5%, 4%) | -1% (-15%, 13%) | -2% (-17%, 12%) | -1% (-3%, 2%) |
| 3-NT |  |  |  |  |  |  |  |  |  |  |
|  | Model 1 | 8% (-1%, 17%) | -3% (-15%, 9%) | 0% (-2%, 2%) | -8% (-24%, 8%) | -10% (-26%, 5%) | -2% (-5%, 1%) | -7% (-16%, 2%) | -8% (-17%, 2%) | -2% (-3%, 0%) |
|  | Model 2 | 8% (0%, 17%) | -4% (-16%, 8%) | 0% (-2%, 2%) | -8% (-24%, 8%) | -11% (-26%, 5%) | -2% (-5%, 1%) | -7% (-16%, 2%) | -8% (-18%, 1%) | -2% (-3%, 0%) |
|  | Model 3 | 9% (0%, 19%) | -2% (-15%, 10%) | 1% (-1%, 3%) | -10% (-27%, 7%) | -14% (-31%, 2%) | -3% (-6%, 0%) | -6% (-16%, 3%) | -8% (-18%, 2%) | -1% (-3%, 1%) |
| NT-proBNP |  |  |  |  |  |  |  |  |  |  |
|  | Model 1 | -2% (-12%, 8%) | -11% (-25%, 2%) | -2% (-4%, 1%) | -7% (-25%, 11%) | -1% (-18%, 16%) | -1% (-4%, 3%) | 0% (-10%, 10%) | 0% (-10%, 11%) | 0% (-2%, 2%) |
|  | Model 2 | -2% (-12%, 8%) | -12% (-25%, 2%) | -2% (-4%, 1%) | -7% (-25%, 11%) | -1% (-18%, 16%) | -1% (-4%, 3%) | 1% (-9%, 11%) | 1% (-10%, 11%) | 0% (-2%, 2%) |
|  | Model 3 | 0% (-9%, 10%) | -7% (-20%, 6%) | -1% (-3%, 1%) | -13% (-31%, 4%) | -8% (-25%, 9%) | -2% (-5%, 1%) | 0% (-10%, 10%) | 2% (-8%, 12%) | 0% (-1%, 2%) |
| Female | | | | | | | | | | |
| PICP |  |  |  |  |  |  |  |  |  |  |
|  | Model 1 | -3% (-12%, 7%) | 1% (-7%, 9%) | 0% (-1%, 2%) | 4% (-10%, 19%) | -2% (-17%, 13%) | 0% (-3%, 3%) | 0% (-5%, 4%) | 5% (-1%, 11%) | 1% (0%, 2%) |
|  | Model 2 | -3% (-12%, 7%) | 1% (-7%, 9%) | 0% (-1%, 2%) | 4% (-10%, 19%) | -2% (-17%, 13%) | 0% (-3%, 3%) | 0% (-5%, 4%) | 5% (-1%, 11%) | 1% (0%, 2%) |
|  | Model 3 | -3% (-12%, 7%) | 1% (-7%, 9%) | 0% (-2%, 2%) | 5% (-10%, 19%) | -2% (-17%, 13%) | 0% (-3%, 3%) | 0% (-5%, 4%) | 5% (-1%, 11%) | 1% (0%, 2%) |
| Hs-TnT |  |  |  |  |  |  |  |  |  |  |
|  | Model 1 | -1% (-8%, 5%) | -2% (-7%, 4%) | 0% (-1%, 1%) | -5% (-15%, 4%) | -8% (-18%, 2%) | -2% (-3%, 0%) | 1% (-1%, 4%) | -1% (-4%, 3%) | 0% (-1%, 1%) |
|  | Model 2 | -1% (-7%, 5%) | -2% (-7%, 4%) | 0% (-1%, 1%) | -5% (-14%, 5%) | -8% (-18%, 2%) | -2% (-3%, 0%) | 1% (-2%, 4%) | -1% (-4%, 3%) | 0% (-1%, 1%) |
|  | Model 3 | -1% (-7%, 5%) | -2% (-7%, 4%) | 0% (-1%, 1%) | -5% (-15%, 5%) | -8% (-18%, 2%) | -2% (-3%, 0%) | 1% (-2%, 4%) | -1% (-4%, 3%) | 0% (-1%, 1%) |
| hsCRP |  |  |  |  |  |  |  |  |  |  |
|  | Model 1 | 0% (-22%, 21%) | 17% (-1%, 36%) | 3% (0%, 6%) | -6% (-39%, 26%) | 16% (-18%, 49%) | 2% (-4%, 8%) | -2% (-12%, 8%) | 3% (-9%, 16%) | 0% (-2%, 2%) |
|  | Model 2 | 2% (-20%, 23%) | 17% (-2%, 35%) | 3% (0%, 6%) | -6% (-39%, 26%) | 13% (-21%, 47%) | 2% (-4%, 7%) | -3% (-13%, 7%) | 3% (-9%, 16%) | 0% (-2%, 2%) |
|  | Model 3 | 1% (-20%, 23%) | 17% (-1%, 36%) | 3% (0%, 6%) | -6% (-38%, 26%) | 14% (-19%, 48%) | 2% (-4%, 8%) | -4% (-14%, 6%) | 2% (-10%, 14%) | 0% (-2%, 2%) |
| 3-NT |  |  |  |  |  |  |  |  |  |  |
|  | Model 1 | -4% (-30%, 21%) | 5% (-17%, 27%) | 1% (-3%, 4%) | -15% (-54%, 24%) | 14% (-26%, 55%) | 1% (-6%, 8%) | -4% (-16%, 8%) | -1% (-16%, 14%) | 0% (-3%, 2%) |
|  | Model 2 | -5% (-30%, 21%) | 4% (-18%, 26%) | 0% (-4%, 4%) | -17% (-57%, 22%) | 14% (-27%, 54%) | 1% (-6%, 8%) | -3% (-15%, 9%) | -1% (-16%, 14%) | 0% (-3%, 2%) |
|  | Model 3 | -3% (-29%, 22%) | 2% (-20%, 25%) | 0% (-4%, 4%) | -15% (-54%, 25%) | 14% (-27%, 55%) | 1% (-6%, 8%) | -3% (-15%, 9%) | -1% (-17%, 14%) | 0% (-3%, 2%) |
| NT-proBNP |  |  |  |  |  |  |  |  |  |  |
|  | Model 1 | 2% (-19%, 23%) | 9% (-10%, 27%) | 2% (-2%, 5%) | -6% (-38%, 27%) | -12% (-46%, 21%) | -2% (-8%, 4%) | -4% (-14%, 6%) | -3% (-15%, 10%) | -1% (-3%, 1%) |
|  | Model 2 | 2% (-19%, 23%) | 9% (-10%, 27%) | 2% (-2%, 5%) | -7% (-40%, 26%) | -13% (-47%, 21%) | -2% (-8%, 4%) | -4% (-14%, 6%) | -2% (-15%, 10%) | -1% (-3%, 1%) |
|  | Model 3 | 3% (-19%, 24%) | 9% (-10%, 28%) | 2% (-2%, 5%) | -9% (-42%, 24%) | -14% (-48%, 20%) | -2% (-8%, 4%) | -5% (-16%, 5%) | -3% (-16%, 10%) | -1% (-3%, 1%) |
| P value for interaction |  |  |  |  |  |  |  |  |  |  |
| PICP |  |  |  |  |  |  |  |  |  |  |
|  | Model 1 | 0.07 | 0.71 | 0.54 | 0.98 | 0.46 | 0.52 | 0.59 | 0.13 | 0.19 |
|  | Model 2 | 0.07 | 0.72 | 0.54 | 0.97 | 0.46 | 0.52 | 0.6 | 0.13 | 0.19 |
|  | Model 3 | 0.08 | 0.87 | 0.67 | 0.98 | 0.47 | 0.53 | 0.64 | 0.21 | 0.29 |
| Hs-TnT |  |  |  |  |  |  |  |  |  |  |
|  | Model 1 | 0.61 | 0.13 | 0.2 | 0.14 | 0.14 | 0.11 | 0.52 | 0.34 | 0.31 |
|  | Model 2 | 0.61 | 0.13 | 0.2 | 0.15 | 0.14 | 0.11 | 0.53 | 0.35 | 0.32 |
|  | Model 3 | 0.56 | 0.17 | 0.25 | 0.26 | 0.24 | 0.2 | 0.84 | 0.58 | 0.55 |
| hsCRP |  |  |  |  |  |  |  |  |  |  |
|  | Model 1 | 0.83 | 0.2 | 0.24 | 0.3 | 0.74 | 0.91 | 0.74 | 0.79 | 0.82 |
|  | Model 2 | 0.83 | 0.21 | 0.25 | 0.3 | 0.76 | 0.93 | 0.74 | 0.78 | 0.81 |
|  | Model 3 | 0.81 | 0.23 | 0.26 | 0.31 | 0.8 | 0.96 | 0.61 | 0.62 | 0.69 |
| 3-NT |  |  |  |  |  |  |  |  |  |  |
|  | Model 1 | 0.21 | 0.57 | 0.78 | 0.82 | 0.16 | 0.22 | 0.62 | 0.91 | 0.99 |
|  | Model 2 | 0.21 | 0.57 | 0.78 | 0.82 | 0.16 | 0.22 | 0.62 | 0.91 | 0.99 |
|  | Model 3 | 0.2 | 0.64 | 0.86 | 0.85 | 0.16 | 0.23 | 0.57 | 0.96 | 0.84 |
| NT-proBNP |  |  |  |  |  |  |  |  |  |  |
|  | Model 1 | 0.58 | 0.04 | 0.24 | 0.67 | 0.99 | 0.91 | 0.29 | 0.8 | 0.82 |
|  | Model 2 | 0.58 | 0.04 | 0.25 | 0.65 | 0.99 | 0.93 | 0.29 | 0.8 | 0.81 |
|  | Model 3 | 0.91 | 0.08 | 0.26 | 0.5 | 0.56 | 0.96 | 0.22 | 0.32 | 0.69 |
| Notes: Mixed models were used. All the concentrations of biomarkers were log-transformed.  Model 1 was adjusted for age, sex; model 2 adjusted for covariates in model 1 + origin, BMI, smoking status, alcohol consumption, and total wearing time; model 3 adjusted for covariates in model 2 + SBP, DBP, HDLc, LDLc, anti-hypertension medication, lipid-lowering medication, insulin, diabetes history. Coefficients (95% confidence intervals) were presented. The coefficients represent the effect of replacing sedentary time with LPA, MVPA, or time in bed on circulating biomarker concentrations at year 5 compared to baseline, and as each year goes by in 5 years. | | | | | | | | | | |

| Supplement Table 4. Association of isotemporal substitution of sedentary time (30min/day) with time in bed and physical activities with change in biomarker between baseline and year 5 by age | | | | | | | | | | |
| --- | --- | --- | --- | --- | --- | --- | --- | --- | --- | --- |
|  |  | LPA | | | MVPA | | | Time in bed | | |
|  |  | Y3 vs. baseline | Y5 vs. baseline | continuous | Y3 vs. baseline | Y5 vs. baseline | continuous | Y3 vs. baseline | Y5 vs. baseline | continuous |
| <65 yrs |  |  |  |  |  |  |  |  |  |  |
| PICP |  |  |  |  |  |  |  |  |  |  |
|  | Model 1 | 0% (-3%, 3%) | 1% (-2%, 4%) | 0% (0%, 1%) | 1% (-3%, 6%) | 0% (-5%, 4%) | 0% (-1%, 1%) | 0% (-2%, 3%) | 0% (-3%, 3%) | 0% (-1%, 1%) |
|  | Model 2 | 0% (-3%, 3%) | 1% (-2%, 4%) | 0% (0%, 1%) | 1% (-3%, 6%) | 0% (-5%, 4%) | 0% (-1%, 1%) | 0% (-2%, 3%) | 0% (-3%, 3%) | 0% (-1%, 1%) |
|  | Model 3 | 0% (-3%, 3%) | 1% (-2%, 4%) | 0% (0%, 1%) | 2% (-3%, 6%) | 0% (-4%, 5%) | 0% (-1%, 1%) | 1% (-2%, 3%) | 0% (-3%, 3%) | 0% (-1%, 1%) |
| Hs-TnT |  |  |  |  |  |  |  |  |  |  |
|  | Model 1 | 0% (-3%, 3%) | -2% (-5%, 1%) | 0% (-1%, 0%) | 1% (-3%, 6%) | -1% (-6%, 3%) | 0% (-1%, 1%) | 1% (-1%, 4%) | -1% (-3%, 2%) | 0% (-1%, 0%) |
|  | Model 2 | 0% (-3%, 3%) | -2% (-5%, 1%) | 0% (-1%, 0%) | 1% (-3%, 6%) | -1% (-6%, 3%) | 0% (-1%, 1%) | 1% (-1%, 4%) | -1% (-3%, 2%) | 0% (-1%, 0%) |
|  | Model 3 | 1% (-2%, 4%) | -2% (-5%, 1%) | 0% (-1%, 0%) | 1% (-4%, 5%) | -2% (-6%, 3%) | 0% (-1%, 1%) | 2% (-1%, 4%) | 0% (-3%, 3%) | 0% (-1%, 1%) |
| hsCRP |  |  |  |  |  |  |  |  |  |  |
|  | Model 1 | -3% (-12%, 5%) | -2% (-10%, 6%) | 0% (-2%, 1%) | -2% (-13%, 10%) | -2% (-14%, 10%) | 0% (-3%, 2%) | -1% (-8%, 6%) | -1% (-9%, 6%) | 0% (-2%, 1%) |
|  | Model 2 | -3% (-12%, 5%) | -2% (-10%, 6%) | 0% (-2%, 1%) | -2% (-14%, 10%) | -2% (-14%, 10%) | 0% (-3%, 2%) | -1% (-8%, 6%) | -2% (-9%, 6%) | 0% (-2%, 1%) |
|  | Model 3 | -4% (-12%, 4%) | -1% (-10%, 7%) | 0% (-2%, 1%) | -1% (-13%, 11%) | -2% (-14%, 10%) | 0% (-3%, 2%) | -2% (-10%, 5%) | -1% (-8%, 7%) | 0% (-2%, 1%) |
| 3-NT |  |  |  |  |  |  |  |  |  |  |
|  | Model 1 | 4% (-3%, 11%) | -5% (-12%, 3%) | -1% (-2%, 1%) | -1% (-12%, 10%) | -4% (-15%, 7%) | -1% (-3%, 1%) | 1% (-6%, 7%) | 0% (-7%, 6%) | 0% (-1%, 1%) |
|  | Model 2 | 4% (-3%, 11%) | -5% (-12%, 3%) | -1% (-2%, 1%) | -1% (-12%, 10%) | -4% (-15%, 7%) | -1% (-3%, 1%) | 1% (-6%, 7%) | 0% (-7%, 6%) | 0% (-1%, 1%) |
|  | Model 3 | 5% (-3%, 12%) | -5% (-13%, 2%) | -1% (-2%, 1%) | -1% (-12%, 9%) | -5% (-16%, 6%) | -1% (-3%, 1%) | 2% (-5%, 8%) | 0% (-7%, 7%) | 0% (-1%, 1%) |
| NT-proBNP |  |  |  |  |  |  |  |  |  |  |
|  | Model 1 | -7% (-15%, 0%) | -7% (-15%, 1%) | -1% (-3%, 0%) | -2% (-14%, 9%) | -4% (-15%, 8%) | -1% (-3%, 2%) | -2% (-8%, 5%) | 0% (-7%, 7%) | 0% (-1%, 1%) |
|  | Model 2 | -7% (-15%, 1%) | -7% (-15%, 1%) | -1% (-3%, 0%) | -2% (-14%, 9%) | -4% (-15%, 8%) | -1% (-3%, 1%) | -1% (-8%, 5%) | 0% (-7%, 7%) | 0% (-1%, 1%) |
|  | Model 3 | -7% (-14%, 1%) | -6% (-14%, 2%) | -1% (-3%, 0%) | -4% (-15%, 8%) | -6% (-17%, 6%) | -1% (-3%, 1%) | -2% (-9%, 5%) | 0% (-7%, 7%) | 0% (-1%, 1%) |
| >=65 yrs |  |  |  |  |  |  |  |  |  |  |
| PICP |  |  |  |  |  |  |  |  |  |  |
|  | Model 1 | -2% (-5%, 1%) | 1% (-1%, 4%) | 0% (0%, 1%) | 0% (-5%, 5%) | -3% (-8%, 2%) | -1% (-2%, 0%) | 0% (-2%, 2%) | 2% (0%, 4%) | 0% (0%, 1%) |
|  | Model 2 | -2% (-5%, 1%) | 2% (-1%, 4%) | 0% (0%, 1%) | 0% (-5%, 5%) | -3% (-8%, 2%) | -1% (-2%, 0%) | 0% (-2%, 2%) | 2% (0%, 4%) | 0% (0%, 1%) |
|  | Model 3 | -2% (-5%, 1%) | 2% (-1%, 5%) | 0% (0%, 1%) | 0% (-5%, 4%) | -3% (-8%, 2%) | -1% (-2%, 0%) | 1% (-1%, 3%) | 2% (0%, 4%) | 0% (0%, 1%) |
| Hs-TnT |  |  |  |  |  |  |  |  |  |  |
|  | Model 1 | -2% (-5%, 1%) | -3% (-6%, 0%) | -1% (-1%, 0%) | 4% (-1%, 9%) | 1% (-4%, 5%) | 0% (-1%, 1%) | -1% (-3%, 1%) | -1% (-3%, 1%) | 0% (-1%, 0%) |
|  | Model 2 | -2% (-5%, 1%) | -3% (-6%, 0%) | -1% (-1%, 0%) | 4% (-1%, 9%) | 0% (-4%, 5%) | 0% (-1%, 1%) | -1% (-3%, 1%) | -1% (-3%, 1%) | 0% (-1%, 0%) |
|  | Model 3 | -2% (-5%, 1%) | -3% (-6%, 0%) | -1% (-1%, 0%) | 4% (-1%, 9%) | 1% (-4%, 5%) | 0% (-1%, 1%) | -1% (-3%, 1%) | -1% (-3%, 1%) | 0% (-1%, 0%) |
| hsCRP |  |  |  |  |  |  |  |  |  |  |
|  | Model 1 | -4% (-15%, 6%) | -6% (-16%, 5%) | -1% (-3%, 1%) | -6% (-24%, 11%) | 5% (-13%, 23%) | 1% (-3%, 4%) | -6% (-13%, 1%) | -2% (-9%, 6%) | 0% (-2%, 1%) |
|  | Model 2 | -4% (-15%, 6%) | -5% (-16%, 5%) | -1% (-3%, 1%) | -6% (-24%, 11%) | 5% (-13%, 23%) | 1% (-3%, 4%) | -6% (-13%, 1%) | -1% (-9%, 6%) | 0% (-2%, 1%) |
|  | Model 3 | -4% (-15%, 6%) | -5% (-16%, 5%) | -1% (-3%, 1%) | -6% (-24%, 11%) | 5% (-13%, 23%) | 1% (-3%, 4%) | -6% (-13%, 1%) | -1% (-9%, 6%) | 0% (-2%, 1%) |
| 3-NT |  |  |  |  |  |  |  |  |  |  |
|  | Model 1 | -6% (-12%, 1%) | -1% (-8%, 6%) | 0% (-2%, 1%) | -13% (-24%, -2%) | -12% (-23%, -1%) | -3% (-5%, 0%) | 0% (-4%, 5%) | 1% (-3%, 6%) | 0% (-1%, 1%) |
|  | Model 2 | -6% (-12%, 1%) | -1% (-8%, 6%) | 0% (-2%, 1%) | -13% (-24%, -2%) | -12% (-23%, -1%) | -3% (-5%, 0%) | 0% (-4%, 5%) | 1% (-3%, 6%) | 0% (-1%, 1%) |
|  | Model 3 | -5% (-12%, 1%) | 0% (-7%, 6%) | 0% (-2%, 1%) | -13% (-24%, -2%) | -11% (-22%, 0%) | -2% (-5%, 0%) | 0% (-5%, 4%) | 1% (-4%, 5%) | 0% (-1%, 1%) |
| NT-proBNP |  |  |  |  |  |  |  |  |  |  |
|  | Model 1 | -1% (-7%, 6%) | -3% (-10%, 3%) | -1% (-2%, 1%) | -5% (-16%, 6%) | -5% (-16%, 6%) | -1% (-3%, 1%) | 1% (-3%, 6%) | 2% (-3%, 6%) | 0% (-1%, 1%) |
|  | Model 2 | -1% (-7%, 6%) | -3% (-10%, 3%) | -1% (-2%, 1%) | -5% (-16%, 6%) | -6% (-17%, 6%) | -1% (-3%, 1%) | 1% (-3%, 6%) | 2% (-3%, 6%) | 0% (-1%, 1%) |
|  | Model 3 | -1% (-8%, 5%) | -3% (-10%, 3%) | -1% (-2%, 1%) | -6% (-17%, 5%) | -5% (-17%, 6%) | -1% (-3%, 1%) | 1% (-3%, 6%) | 1% (-3%, 6%) | 0% (-1%, 1%) |
| P value for interaction |  |  |  |  |  |  |  |  |  |  |
| PICP |  |  |  |  |  |  |  |  |  |  |
|  | Model 1 | 0.39 | 0.03 | 0.07 | 0.49 | 0.92 | 0.89 | 0.77 | 0.09 | 0.13 |
|  | Model 2 | 0.39 | 0.03 | 0.07 | 0.5 | 0.92 | 0.89 | 0.77 | 0.08 | 0.13 |
|  | Model 3 | 0.4 | 0.01 | 0.03 | 0.49 | 0.99 | 0.96 | 0.8 | 0.1 | 0.15 |
| Hs-TnT |  |  |  |  |  |  |  |  |  |  |
|  | Model 1 | 0.32 | 0.11 | 0.19 | 0.22 | 0.57 | 0.73 | 0.67 | 0.51 | 0.49 |
|  | Model 2 | 0.32 | 0.11 | 0.73 | 0.23 | 0.57 | 0.2 | 0.68 | 0.51 | 0.5 |
|  | Model 3 | 0.21 | 0.19 | 0.08 | 0.08 | 0.75 | 0.97 | 0.79 | 0.69 | 0.58 |
| hsCRP |  |  |  |  |  |  |  |  |  |  |
|  | Model 1 | 0.86 | 0.24 | 0.28 | 0.93 | 0.4 | 0.44 | 0.42 | 0.53 | 0.49 |
|  | Model 2 | 0.86 | 0.24 | 0.28 | 0.93 | 0.41 | 0.45 | 0.42 | 0.54 | 0.5 |
|  | Model 3 | 0.78 | 0.22 | 0.8 | 0.87 | 0.37 | 0.56 | 0.43 | 0.37 | 0.38 |
| 3-NT |  |  |  |  |  |  |  |  |  |  |
|  | Model 1 | 0.38 | 0.84 | 0.69 | 0.43 | 0.37 | 0.76 | 0.11 | 0.47 | 0.12 |
|  | Model 2 | 0.11 | 0.47 | 0.69 | 0.38 | 0.83 | 0.75 | 0.08 | 0.18 | 0.12 |
|  | Model 3 | 0.08 | 0.59 | 0.63 | 0.43 | 0.91 | 0.23 | 0.04 | 0.16 | 0.84 |
| NT-proBNP |  |  |  |  |  |  |  |  |  |  |
|  | Model 1 | 0.32 | 0.93 | 0.36 | 0.98 | 0.32 | 0.56 | 1 | 0.88 | 0.86 |
|  | Model 2 | 0.99 | 0.32 | 0.36 | 0.92 | 0.53 | 0.55 | 0.99 | 0.88 | 0.86 |
|  | Model 3 | 0.38 | 0.96 | 0.61 | 0.9 | 0.64 | 0.17 | 0.93 | 0.59 | 0.83 |
| Notes: Mixed models were used. All the concentrations of biomarkers were log-transformed.  Model 1 was adjusted for age, sex; model 2 adjusted for covariates in model 1 + origin, BMI, smoking status, alcohol consumption, and total wearing time; model 3 adjusted for covariates in model 2 + SBP, DBP, HDLc, LDLc, anti-hypertension medication, lipid-lowering medication, insulin, diabetes history. Coefficients (95% confidence intervals) were presented. The coefficients represent the effect of replacing sedentary time with LPA, MVPA, or time in bed on circulating biomarker concentrations at year 5 compared to baseline, and as each year goes by in 5 years. | | | | | | | | | | |

| Supplemental Table 5. Association of isotemporal substitution of visit 3 sedentary time (30min/day) with time in bed and physical activities with change in biomarkers between year 3 and year 5 by sex | | | | | | | |
| --- | --- | --- | --- | --- | --- | --- | --- |
|  |  | LPA | | MVPA | | Time in bed | |
|  |  | Y5 vs. Y3 | continuous | Y5 vs. Y3 | continuous | Y5 vs. Y3 | continuous |
| Male |  |  |  |  |  |  |  |
| PICP |  |  |  |  |  |  |  |
|  | Model 1 | 3% (-4%, 10%) | 1% (-2%, 5%) | -4% (-13%, 4%) | -2% (-6%, 2%) | -2% (-8%, 3%) | -1% (-4%, 2%) |
|  | Model 2 | 3% (-4%, 10%) | 2% (-2%, 5%) | -5% (-13%, 3%) | -2% (-7%, 2%) | -2% (-8%, 4%) | -1% (-4%, 2%) |
|  | Model 3 | 1% (-6%, 8%) | 0% (-3%, 4%) | -4% (-12%, 5%) | -2% (-6%, 2%) | -2% (-8%, 4%) | -1% (-4%, 2%) |
| Hs-TnT |  |  |  |  |  |  |  |
|  | Model 1 | -1% (-6%, 4%) | 0% (-3%, 2%) | -1% (-7%, 5%) | 0% (-3%, 3%) | 1% (-2%, 5%) | 1% (-1%, 3%) |
|  | Model 2 | -1% (-6%, 4%) | -1% (-3%, 2%) | -1% (-7%, 5%) | 0% (-3%, 3%) | 1% (-3%, 5%) | 1% (-1%, 3%) |
|  | Model 3 | -1% (-6%, 4%) | 0% (-3%, 2%) | 1% (-5%, 7%) | 1% (-2%, 4%) | 1% (-2%, 5%) | 1% (-1%, 3%) |
| hsCRP |  |  |  |  |  |  |  |
|  | Model 1 | 11% (-7%, 29%) | 6% (-4%, 15%) | 5% (-17%, 28%) | 3% (-8%, 14%) | -2% (-17%, 13%) | -1% (-8%, 6%) |
|  | Model 2 | 9% (-9%, 27%) | 4% (-5%, 13%) | 8% (-14%, 31%) | 4% (-7%, 15%) | -3% (-18%, 11%) | -2% (-9%, 6%) |
|  | Model 3 | 10% (-10%, 30%) | 5% (-5%, 15%) | 4% (-20%, 28%) | 2% (-10%, 14%) | 0% (-16%, 15%) | 0% (-8%, 8%) |
| 3-NT |  |  |  |  |  |  |  |
|  | Model 1 | -9% (-25%, 6%) | -5% (-12%, 3%) | -2% (-21%, 17%) | -1% (-11%, 8%) | -10% (-22%, 3%) | -5% (-11%, 1%) |
|  | Model 2 | -10% (-26%, 5%) | -5% (-13%, 3%) | -2% (-22%, 17%) | -1% (-11%, 8%) | -10% (-23%, 2%) | -5% (-11%, 1%) |
|  | Model 3 | -7% (-24%, 10%) | -4% (-12%, 5%) | -5% (-26%, 16%) | -2% (-13%, 8%) | -8% (-22%, 5%) | -4% (-11%, 3%) |
| NT-proBNP |  |  |  |  |  |  |  |
|  | Model 1 | -2% (-14%, 11%) | -1% (-7%, 5%) | -6% (-21%, 10%) | -3% (-11%, 5%) | -5% (-15%, 5%) | -3% (-8%, 2%) |
|  | Model 2 | -2% (-14%, 11%) | -1% (-7%, 5%) | -6% (-22%, 10%) | -3% (-11%, 5%) | -5% (-15%, 5%) | -3% (-8%, 2%) |
|  | Model 3 | -3% (-16%, 9%) | -2% (-8%, 4%) | -4% (-19%, 11%) | -2% (-9%, 5%) | -5% (-14%, 5%) | -2% (-7%, 2%) |
| Female |  |  |  |  |  |  |  |
| PICP |  |  |  |  |  |  |  |
|  | Model 1 | -5% (-17%,8%) | -2% (-8%,4%) | -2% (-18%,14%) | -1% (-9%,7%) | 0% (-7%,7%) | 0% (-4%,4%) |
|  | Model 2 | -2% (-15%,11%) | -1% (-7%,5%) | 1% (-16%,17%) | 0% (-8%,9%) | 2% (-6%,9%) | 1% (-3%,5%) |
|  | Model 3 | 0% (-14%,14%) | 0% (-7%,7%) | 0% (-18%,17%) | 0% (-9%,9%) | 2% (-7%,10%) | 1% (-4%,5%) |
| Hs-TnT |  |  |  |  |  |  |  |
|  | Model 1 | -2% (-11%,6%) | -1% (-5%,3%) | -6% (-16%,5%) | -3% (-8%,3%) | 0% (-5%,6%) | 0% (-3%,3%) |
|  | Model 2 | -2% (-11%,6%) | -1% (-5%,3%) | -6% (-17%,5%) | -3% (-9%,2%) | 0% (-5%,6%) | 0% (-3%,3%) |
|  | Model 3 | -1% (-10%,8%) | -1% (-5%,4%) | -4% (-15%,7%) | -2% (-7%,4%) | 2% (-3%,8%) | 1% (-2%,4%) |
| hsCRP |  |  |  |  |  |  |  |
|  | Model 1 | 4% (-19%,27%) | 2% (-10%,14%) | -1% (-31%,29%) | -1% (-16%,14%) | 0% (-14%,14%) | 0% (-7%,7%) |
|  | Model 2 | 2% (-21%,25%) | 1% (-11%,12%) | -3% (-33%,27%) | -1% (-16%,14%) | 1% (-13%,15%) | 1% (-6%,8%) |
|  | Model 3 | 1% (-23%,25%) | 0% (-12%,12%) | -9% (-39%,22%) | -4% (-20%,11%) | -4% (-20%,12%) | -2% (-10%,6%) |
| 3-NT |  |  |  |  |  |  |  |
|  | Model 1 | -4% (-16%,9%) | -2% (-8%,5%) | 10% (-6%,26%) | 5% (-3%,13%) | 0% (-8%,9%) | 0% (-4%,5%) |
|  | Model 2 | -4% (-17%,8%) | -2% (-9%,4%) | 9% (-7%,25%) | 4% (-4%,13%) | 0% (-9%,8%) | 0% (-4%,4%) |
|  | Model 3 | -5% (-18%,8%) | -2% (-9%,4%) | 8% (-9%,24%) | 4% (-4%,12%) | -1% (-10%,8%) | -1% (-5%,4%) |
| NT-proBNP |  |  |  |  |  |  |  |
|  | Model 1 | 5% (-23%,33%) | 3% (-11%,17%) | -1% (-37%,35%) | -1% (-19%,18%) | 3% (-15%,20%) | 1% (-7%,10%) |
|  | Model 2 | 7% (-23%,36%) | 3% (-11%,18%) | 0% (-38%,37%) | 0% (-19%,19%) | 4% (-14%,23%) | 2% (-7%,11%) |
|  | Model 3 | 6% (-24%,35%) | 3% (-12%,18%) | 3% (-35%,41%) | 1% (-17%,20%) | 6% (-13%,25%) | 3% (-6%,13%) |
| P value for interaction |  |  |  |  |  |  |  |
| PICP |  |  |  |  |  |  |  |
|  | Model 1 | 0.69 | 0.69 | 0.94 | 0.94 | 0.05 | 0.05 |
|  | Model 2 | 0.89 | 0.89 | 0.72 | 0.72 | 0.03 | 0.03 |
|  | Model 3 | 0.6 | 0.6 | 0.65 | 0.65 | 0.08 | 0.08 |
| Hs-TnT |  |  |  |  |  |  |  |
|  | Model 1 | 0.24 | 0.24 | 0.72 | 0.72 | 0.48 | 0.48 |
|  | Model 2 | 0.54 | 0.54 | 0.83 | 0.83 | 0.62 | 0.62 |
|  | Model 3 | 0.38 | 0.38 | 0.94 | 0.94 | 0.23 | 0.23 |
| hsCRP |  |  |  |  |  |  |  |
|  | Model 1 | 0.29 | 0.29 | 0.54 | 0.54 | 0.18 | 0.18 |
|  | Model 2 | 0.98 | 0.98 | 0.75 | 0.75 | 0.27 | 0.27 |
|  | Model 3 | 0.86 | 0.86 | 0.89 | 0.89 | 0.27 | 0.27 |
| 3-NT |  |  |  |  |  |  |  |
|  | Model 1 | 0.05 | 0.05 | 0.04 | 0.04 | 0.3 | 0.3 |
|  | Model 2 | 0.08 | 0.08 | 0.06 | 0.06 | 0.29 | 0.29 |
|  | Model 3 | 0.12 | 0.12 | 0.09 | 0.09 | 0.36 | 0.36 |
| NT-proBNP |  |  |  |  |  |  |  |
|  | Model 1 | 0.55 | 0.55 | 0.42 | 0.46 | 0.92 | 0.92 |
|  | Model 2 | 0.6 | 0.6 | 0.43 | 0.43 | 0.95 | 0.95 |
|  | Model 3 | 0.31 | 0.31 | 0.69 | 0.69 | 0.45 | 0.45 |
| Notes: Mixed models were used. All the concentrations of biomarkers were log-transformed.  Model 1 was adjusted for age, sex, intervention group; model 2 adjusted for covariates in model 1 + origin, BMI, smoking status, alcohol consumption, and total wearing time; model 3 adjusted for covariates in model 2 + SBP, DBP, HDLc, LDLc, anti-hypertension medication, lipid-lowering medication, insulin, diabetes history. Coefficients (95% confidence intervals) were presented. The coefficients represent the effect of replacing sedentary time with LPA, MVPA, or time in bed on circulating biomarker concentrations at year 5 compared to year 3, and as each year goes by in 3 years. | | | | | | | |

| Supplemental Table 6. Association of isotemporal substitution of visit 3 sedentary time (30min/day) with time in bed and physical activities with change in biomarkers between year 3 and year 5 by age | | | | | | | |
| --- | --- | --- | --- | --- | --- | --- | --- |
|  |  | LPA | | MVPA | | Time in bed | |
|  |  | Y5 vs. Y3 | continuous | Y5 vs. Y3 | continuous | Y5 vs. Y3 | continuous |
| <65 yrs |  |  |  |  |  |  |  |
| PICP |  |  |  |  |  |  |  |
|  | Model 1 | -2% (-11%, 7%) | -1% (-5%, 4%) | -8% (-20%, 4%) | -4% (-10%, 2%) | -3% (-9%, 4%) | -1% (-5%, 2%) |
|  | Model 2 | -1% (-10%, 7%) | -1% (-5%, 4%) | -8% (-21%, 4%) | -4% (-10%, 2%) | -3% (-10%, 4%) | -1% (-5%, 2%) |
|  | Model 3 | -6% (-15%, 4%) | -3% (-8%, 2%) | -8% (-20%, 5%) | -4% (-10%, 3%) | -4% (-11%, 3%) | -2% (-5%, 1%) |
| Hs-TnT |  |  |  |  |  |  |  |
|  | Model 1 | -4% (-9%, 2%) | -2% (-5%, 1%) | -2% (-10%, 6%) | -1% (-5%, 3%) | 1% (-3%, 5%) | 0% (-2%, 3%) |
|  | Model 2 | -4% (-10%, 2%) | -2% (-5%, 1%) | -1% (-10%, 7%) | -1% (-5%, 3%) | 1% (-3%, 5%) | 1% (-2%, 3%) |
|  | Model 3 | -4% (-10%, 2%) | -2% (-5%, 1%) | 3% (-5%, 11%) | 2% (-3%, 6%) | 2% (-2%, 6%) | 1% (-1%, 3%) |
| hsCRP |  |  |  |  |  |  |  |
|  | Model 1 | 17% (-3%, 37%) | 8% (-1%, 18%) | -9% (-36%, 19%) | -4% (-18%, 9%) | -2% (-16%, 12%) | -1% (-8%, 6%) |
|  | Model 2 | 13% (-6%, 33%) | 7% (-3%, 17%) | -1% (-28%, 26%) | 0% (-14%, 13%) | -2% (-16%, 13%) | -1% (-8%, 7%) |
|  | Model 3 | 14% (-8%, 36%) | 7% (-4%, 18%) | -6% (-36%, 23%) | -3% (-18%, 12%) | 3% (-13%, 19%) | 2% (-6%, 10%) |
| 3-NT |  |  |  |  |  |  |  |
|  | Model 1 | -14% (-31%, 4%) | -7% (-16%, 2%) | 8% (-17%, 32%) | 4% (-8%, 16%) | -14% (-26%, -1%) | -7% (-13%, 0%) |
|  | Model 2 | -13% (-30%, 5%) | -6% (-15%, 2%) | 7% (-17%, 32%) | 4% (-9%, 16%) | -14% (-27%, -2%) | -7% (-14%, -1%) |
|  | Model 3 | -12% (-32%, 8%) | -6% (-16%, 4%) | 7% (-20%, 35%) | 4% (-10%, 17%) | -15% (-29%, -1%) | -8% (-15%, 0%) |
| NT-proBNP |  |  |  |  |  |  |  |
|  | Model 1 | 2% (-14%, 18%) | 1% (-7%, 9%) | -4% (-26%, 18%) | -2% (-13%, 9%) | -3% (-15%, 9%) | -2% (-8%, 4%) |
|  | Model 2 | 2% (-15%, 18%) | 1% (-7%, 9%) | -4% (-26%, 19%) | -2% (-13%, 9%) | -3% (-15%, 9%) | -2% (-8%, 4%) |
|  | Model 3 | -3% (-19%, 14%) | -1% (-10%, 7%) | 3% (-19%, 26%) | 2% (-10%, 13%) | -1% (-13%, 11%) | -1% (-6%, 5%) |
| >= 65 yrs |  |  |  |  |  |  |  |
| PICP |  |  |  |  |  |  |  |
|  | Model 1 | 5% (-2%,13%) | 3% (-1%,6%) | -1% (-10%,8%) | 0% (-5%,4%) | 0% (-5%,6%) | 0% (-2%,3%) |
|  | Model 2 | 7% (-1%,15%) | 4% (0%,7%) | 0% (-9%,8%) | 0% (-5%,4%) | 1% (-4%,7%) | 1% (-2%,3%) |
|  | Model 3 | 7% (-1%,15%) | 4% (0%,8%) | 1% (-8%,10%) | 1% (-4%,5%) | 1% (-4%,6%) | 0% (-2%,3%) |
| Hs-TnT |  |  |  |  |  |  |  |
|  | Model 1 | 2% (-3%,7%) | 1% (-1%,4%) | -2% (-7%,4%) | -1% (-4%,2%) | 1% (-2%,5%) | 1% (-1%,2%) |
|  | Model 2 | 2% (-3%,7%) | 1% (-1%,4%) | -2% (-7%,4%) | -1% (-4%,2%) | 1% (-2%,5%) | 1% (-1%,2%) |
|  | Model 3 | 2% (-3%,7%) | 1% (-1%,4%) | -2% (-7%,4%) | -1% (-4%,2%) | 1% (-2%,5%) | 1% (-1%,2%) |
| hsCRP |  |  |  |  |  |  |  |
|  | Model 1 | 8% (-12%,29%) | 4% (-6%,15%) | 15% (-9%,39%) | 7% (-5%,19%) | 1% (-13%,16%) | 1% (-7%,8%) |
|  | Model 2 | 6% (-15%,27%) | 3% (-8%,13%) | 14% (-10%,38%) | 7% (-5%,19%) | 0% (-14%,15%) | 0% (-7%,7%) |
|  | Model 3 | 6% (-16%,28%) | 3% (-8%,14%) | 13% (-12%,37%) | 6% (-6%,19%) | 1% (-14%,15%) | 0% (-7%,8%) |
| 3-NT |  |  |  |  |  |  |  |
|  | Model 1 | -2% (-16%,12%) | -1% (-8%,6%) | -5% (-21%,11%) | -2% (-10%,5%) | 1% (-9%,10%) | 0% (-5%,5%) |
|  | Model 2 | -3% (-18%,11%) | -2% (-9%,5%) | -6% (-21%,10%) | -3% (-11%,5%) | 0% (-10%,10%) | 0% (-5%,5%) |
|  | Model 3 | -3% (-18%,11%) | -2% (-9%,5%) | -7% (-23%,9%) | -3% (-11%,5%) | 0% (-10%,10%) | 0% (-5%,5%) |
| NT-proBNP |  |  |  |  |  |  |  |
|  | Model 1 | 3% (-12%,18%) | 2% (-6%,9%) | -9% (-26%,8%) | -4% (-13%,4%) | 3% (-7%,14%) | 2% (-4%,7%) |
|  | Model 2 | 4% (-11%,20%) | 2% (-6%,10%) | -9% (-27%,8%) | -5% (-13%,4%) | 4% (-7%,14%) | 2% (-3%,7%) |
|  | Model 3 | 5% (-11%,20%) | 2% (-5%,10%) | -11% (-28%,6%) | -6% (-14%,3%) | 4% (-6%,15%) | 2% (-3%,8%) |
| P value for interaction |  |  |  |  |  |  |  |
| PICP |  |  |  |  |  |  |  |
|  | Model 1 | 0.81 | 0.02 | 0.06 | 0.73 | 0.16 | 0.2 |
|  | Model 2 | 0.008 | 0.008 | 0.8 | 0.97 | 0.11 | 0.14 |
|  | Model 3 | 0.005 | 0.006 | 0.82 | 0.98 | 0.11 | 0.13 |
| Hs-TnT |  |  |  |  |  |  |  |
|  | Model 1 | 0.11 | 0.14 | 0.85 | 0.73 | 0.46 | 0.39 |
|  | Model 2 | 0.13 | 0.13 | 0.93 | 0.88 | 0.54 | 0.45 |
|  | Model 3 | 0.11 | 0.1 | 0.56 | 0.73 | 0.37 | 0.33 |
| hsCRP |  |  |  |  |  |  |  |
|  | Model 1 | 0.86 | 0.86 | 0.62 | 0.59 | 0.68 | 0.71 |
|  | Model 2 | 0.97 | 0.95 | 0.96 | 0.97 | 0.51 | 0.45 |
|  | Model 3 | 0.77 | 0.74 | 0.72 | 0.79 | 0.51 | 0.46 |
| 3-NT |  |  |  |  |  |  |  |
|  | Model 1 | 0.29 | 0.29 | 0.82 | 0.8 | 0.33 | 0.32 |
|  | Model 2 | 0.36 | 0.36 | 0.96 | 0.94 | 0.39 | 0.38 |
|  | Model 3 | 0.44 | 0.44 | 0.92 | 0.95 | 0.29 | 0.28 |
| NT-proBNP |  |  |  |  |  |  |  |
|  | Model 1 | 0.56 | 0.55 | 0.82 | 0.86 | 0.45 | 0.41 |
|  | Model 2 | 0.63 | 0.65 | 0.69 | 0.75 | 0.69 | 0.43 |
|  | Model 3 | 0.71 | 0.72 | 0.56 | 0.59 | 0.36 | 0.34 |
| Notes: Mixed models were used. All the concentrations of biomarkers were log-transformed.  Model 1 was adjusted for age, sex, intervention group; model 2 adjusted for covariates in model 1 + origin, BMI, smoking status, alcohol consumption, and total wearing time; model 3 adjusted for covariates in model 2 + SBP, DBP, HDLc, LDLc, anti-hypertension medication, lipid-lowering medication, insulin, diabetes history. Coefficients (95% confidence intervals) were presented. The coefficients represent the effect of replacing sedentary time with LPA, MVPA, or time in bed on circulating biomarker concentrations at year 5 compared to year 3, and as each year goes by in 3 years. | | | | | | | |
